# Cigarette Smoking Across Life from 1946 to 2018: Harmonisation of Four British Birth Cohort Studies

**DOI:** 10.1101/2024.12.06.24318606

**Authors:** Liam Wright, Loren Kock, Harry Tattan-Birch, David Bann

**Affiliations:** Centre for Longitudinal Studies, Social Research Institute, University College London; Department of Behavioural Science and Health, University College London

**Keywords:** cross-cohort analysis, life course analysis, cohort effects, smoking, tobacco

## Abstract

**Background and Aims:** Tobacco smoking has declined dramatically in the many high-income countries over the past seventy years. Studies that have mapped this trend have relied on repeat cross-sectional or retrospectively measured smoking data which has limitations regarding accurate measurement, inclusion of early smokers, and capturing of within-person change over time. Here, we (1) introduce a new resource detailing harmonisable smoking data in four British birth cohort studies spanning 1946-2018 and (2) use this data to document age and cohort changes in smoking.

**Methods:** We used prospectively and longitudinally measured smoking data in four British Birth Cohort Studies, born 1946, 1958, 1970 and 2000/02, respectively, to examine changes in the prevalence of daily smoking and cigarettes smoked per day between cohorts and within cohorts over the life course in males and females combined and stratified by sex.

**Results:** The prevalence of smoking and the average number of cigarettes smoked by daily smokers declined between each successive cohort. Males smoked more and with greater intensity than females, on average, though sex differences were smaller in latter cohorts. Within a cohort, the prevalence and intensity of smoking peaked in early adulthood (< age 30y) and declined thereafter; participants who continued to smoke daily, smoked fewer cigarettes as they grew older.

**Conclusion:** **S**moking prevalence and cigarette consumption declined substantially between cohorts born across the latter half of the twentieth and early twenty-first centuries. The British Birth Cohorts represent a unique and largely underutilized resource for investigating trends in smoking across life (prenatal to old age) and by year of birth (1946-2001), including changes in the determinants, correlates, and consequences of smoking. We provide syntax and information on items on smoking in these cohorts to catalyse future research, also available at: <u>osf.io/54w6q</u>.

## Introduction

The British Birth Cohort Studies have been described as ‘jewels in the crown’ of UK social science (Pearson, 2016). These studies, which follow nationally representative samples of individuals born in 1946, 1958, 1970 and 2000/02, respectively, offer a unique window into the major social changes that have occurred in Great Britain since the Second World War, such as the expansion of higher education, growing female labour force participation, changes in social mobility, and increasing obesity rates (Anders & Dorsett, 2017; Ferri et al., 2003; Gregg & Wadsworth, 2011; Johnson et al., 2015). Beyond enabling researchers to document the size and timing of these trends, these cohorts allow investigation into changes in the determinants – and consequences – of socioeconomic and health outcomes, both between generations and over the life course (Bann et al., 2018, 2023; Sellers et al., 2019; Wright et al., 2024).

One health behaviour that has undergone substantial change over the past seventy years is cigarette smoking. The prevalence of smoking, which reached 80% among men in Great Britain in 1950 (Peto et al., 2000), declined dramatically following the discovery of tobacco’s role in the aetiology of lung cancer and subsequent implementation of taxes, bans and other measures intended to curtail its use (Beard et al., 2019; Christopoulou, 2015). Approximately 12% of UK adults now smoke (Buss et al., 2024; Office for National Statistics, 2024), one of the lowest rates in Western Europe (Ritchie & Roser, 2023), though smoking remains a leading cause of preventable death and social inequalities in health (Harker, 2023; Payne et al., 2022; Vaccarella et al., 2023).

Cross-sectionally, younger adults are more likely to smoke cigarettes as initiation typically occurs during adolescence or early adulthood, with more individuals quitting than taking up smoking thereafter (Christopoulou, 2015; Office for National Statistics, 2024). Yet, these age differences belie large cohort effects: at a given age, individuals born more recently are much less likely to have been a regular smoker (Christopoulou, 2015; Davy, 2006; Kemm, 2001; Opazo Breton et al., 2022). The precise pattern differs by sex (Christopoulou, 2015; Kemm, 2001): smoking prevalence among women peaked later than among men (though at a lower level) and exhibited a slower decline over time (Christopoulou, 2015).

Existing studies on cross-cohort changes in cigarette consumption have used repeat cross-sectional data to create pseudo-cohorts or have relied upon (often older) people reporting their smoking history retrospectively (Davy, 2006; Holford et al., 2014; Kemm, 2001; Lillard & Christopoulou, 2015; Marcon et al., 2018; Opazo Breton et al., 2022). In contrast, smoking in each of the British Birth Cohort Studies has been measured prospectively and on multiple occasions over the life course. This has several advantages over the other approaches. Importantly, prospective measurement avoids survivor and recall biases and tracking the same individuals across time allows individual trajectories to be examined and complex behaviours (e.g., relapse or changes in smoking intensity) to be defined. Some early smokers will not appear in retrospectively measured or repeat cross-sectional data due to premature mortality and earlier smoking may alternatively be misremembered or not reported, including – feasibly – if smoking is stigmatized, which is increasingly the case (Gallup Inc., 2007). Further, repeated cross-sectional data, in particular, cannot distinguish between changes in smoking being due to differences in uptake, cessation or premature mortality (Opazo Breton et al., 2022) and cannot clarify whether changes in smoking intensity are due to *within person* changes in consumption or differences in quit rates according to amount smoked, limiting the lessons that can be drawn.

Beyond avoiding these problems, birth cohort studies expand the range of research questions that can be asked. Most saliently, they enable investigation into life course determinants and consequences of smoking – for instance, on associations between maternal smoking and adult health (Power et al., 2010). The British Birth Cohort Studies, in particular, represent a rich seam of information for researchers to use. Each contains thousands of participants and tens of thousands of variables and includes links with administrative data, biological assessments and genetic assays, much of which can be harmonized across the studies (O’Neill et al., 2019; Shireby et al., 2024) and used to perform appropriately powered analyses of individual trajectories and cross-cohort change, including for important sub-groups such as gender and socioeconomic groups (Bann et al., 2018).

In this study, we present the output of an effort to collate and harmonise data on smoking behaviour from the four British Birth Cohort Studies. We use this data to examine cohort and age-related changes in several measures of tobacco consumption – including current and former daily smoking and smoking intensity (cigarettes per day) – overall and separately by gender, leveraging prospective and repeated measurement of participants. We reasoned that increased knowledge of smoking’s harms and the implementation of measures (e.g., taxes) to curtail tobacco’s use would mean that individuals have become less likely to start smoking, more likely to quit, and more likely to reduce the number of cigarettes they consume either to reduce health risks or lessen the financial cost of smoking (e.g., by drawing more nicotine from each cigarette; Adda & Cornaglia, 2006). Therefore, our hypotheses were that: population prevalence of daily or ever smoking and the average number cigarettes smoked by smokers would be progressively lower *between* cohorts, and *within* a cohort, the prevalence of daily smoking among the total population and the number of cigarettes smoked daily by ‘sustained’ smokers would be highest in early adulthood (∼age 25y), declining thereafter.

Accompanying this paper is code to generate the harmonized variables so that researchers can use the data in their own analyses. We also provide detailed information on the smoking-related data available in each cohort as a reference source (see Supplementary Information and https://cls-data.github.io/smoking-in-the-cohorts/).

## Methods

### Participants

The MRC National Survey of Health and Development (hereafter, 1946c) follows a sample of individuals born in mainland Britain (England, Scotland, or Wales) during a single week of March 1946. Participants were recruited by sampling singleton births, with individuals from households with non-manual employment oversampled. The National Child Development Study (hereafter, 1958c) and the 1970 British Cohort Study (hereafter, 1970c) track all individuals born in mainland Britain in single weeks of March 1958 and April 1970, respectively. Immigrants to the UK born in these same weeks were later added using school enrollment information. The Millennium Cohort Study (hereafter, 2001c) follows a sample of individuals from across the UK born between 2000/02. Participants were recruited using a two-stage stratified sampling design with participants sampled from selected postcode areas. Individuals from Northern Ireland, Scotland and Wales, ethnic minority backgrounds, or disadvantaged areas were oversampled. Given differences in cohort eligibility and increasing ethnic diversity within the UK, to increase comparability, we restricted our analysis to singletons of White ethnicity born in England, Scotland, or Wales.

Each cohort and survey sweep has received ethical approval and obtained appropriate consents according to guidance in place at data collection. Further details on each study is available in cohort profiles (Connelly & Platt, 2014; Elliott & Shepherd, 2006; Kuh et al., 2011; Power & Elliott, 2006; Sullivan et al., 2023; Wadsworth et al., 2006).

### Cigarette Smoking

Data on smoking has been collected as various points over participants’ life courses, including during adolescence for the 1958c, 1970c and 2001c, as well as on participants themselves and their family members (e.g., parents) and, more recently, on e-cigarette use. The ages at which follow-up sweeps have been performed differs across studies, but there is overlap between cohorts at several ages, including age 42/43y when the 1946c, 1958c and 1970c were each interviewed (1989, 2000/01, and 2012, respectively). A full description of the smoking data available in each cohort is provided in the Supplementary Information (as the cohorts are ongoing, a live version of the document is also available at https://osf.io/54w6q/). Here we use variables for current daily smoking (participants and participants’ mothers and fathers), smoking intensity (cigarettes per day; participants only), ever and former daily smoking (participants only), and daily smoking during pregnancy (participants’ mothers). Data on smoking intensity was not available in the 2001c and data on smoking during pregnancy was not available in the 1946c.

### Statistical Analysis

We calculated descriptive statistics for each outcome, separately for each cohort sweep, for males and females combined and stratified by sex. For binary variables (e.g., current daily smoking), we calculated the proportion reporting the behaviour. For smoking intensity, we calculated the mean and standard deviation for (a) the total population, (b) daily smokers (at a given sweep), and (c) sets of ‘sustained’ smokers who reported daily smoking at consecutive sweeps (e.g., at each sweep between age 20y to 43y in the 1946c). Focusing on changes in smoking intensity among sets of sustained smokers allowed us to account for compositional issues whereby individuals with the greatest nicotine dependence may quit less and therefore make up a progressively larger share of smokers over time.

As each cohort was subject to missingness over time, to address the loss of power and potential bias caused by attrition (Silverwood et al., 2024), we used multiple imputation (MI) to impute missing values (m = 40, iterations = 10). Data were inputted in wide format so that earlier and later measurements of smoking behaviour could be used as predictors in imputation models. Observations were removed post-imputation where the participant was known to be dead at that sweep. We used Classification and Regression Trees (CART), a machine learning method, to account for potentially non-linearity and collinearity in the relationship between variables. We included several earlier life predictors as auxiliary variables in imputation models: sex, participants’ verbal cognitive ability at age 10/11y, country of birth, parental socioeconomic position (family social class and mother’s and father’s education level), age, and parental height and body mass index [kg/m^2^]). As a robustness check, we instead used inverse probability weights (IPW) to account for attrition with weights generated using these same auxiliary variables plus measurements of parental smoking, given these were collected in childhood; remaining item-missingness for the variables in the response models was accounted for using MI.

Both weighting and MI assume that data are ‘missing at random’ (i.e., missingness is independent of the true value conditional on imputation model covariates). This may be incorrect if smokers are more likely to drop-out of the study (though, note, earlier smoking observations are included in our imputation models). To explore the sensitivity of our results to selective attrition, we repeated (IPW) analyses for participants’ prevalence of daily smoking assuming that smokers who dropped out of the study continued to smoke following the ‘Russell Standard’ (West et al., 2005).

We carried out all analyses using R version 4.3.1 (R Core Team, 2023). Given the 1946c and 2001c used stratified study designs, we accounted for complex survey design using study-specific probability (recruitment) weights.

## Results

### Cohort Members’ Cigarette Smoking

There were 5,362 eligible participants in the 1946c, 16,178 in the 1958c, 16,036 in the 1970c, and 13,366 in the 2001c. There were sizeable cross-cohort differences in the proportion of participants who smoked daily (top left panel, Figure 1). At age 16/17y, the prevalence of daily smoking was 27.7% (26.9%, 28.5%) in the 1958c, 19.8% (18.9%, 20.7%) in the 1970c, and 10.4% (9.5%, 11.2%) in the 2001c. Figures at age 42/43y were 33.6% (31.8%, 35.5%) in the 1946c, 27.3% (26.5%, 28.2%) in the 1958c, and 22.1% (21.3%, 22.8%) in the 1970c. Prevalence increased during adolescence, reached its peak in early adulthood (ages 20-30y) and declined thereafter. Age related declines during adulthood were similar in the 1946c, 1958, and 1970c.

**Figure 1:**
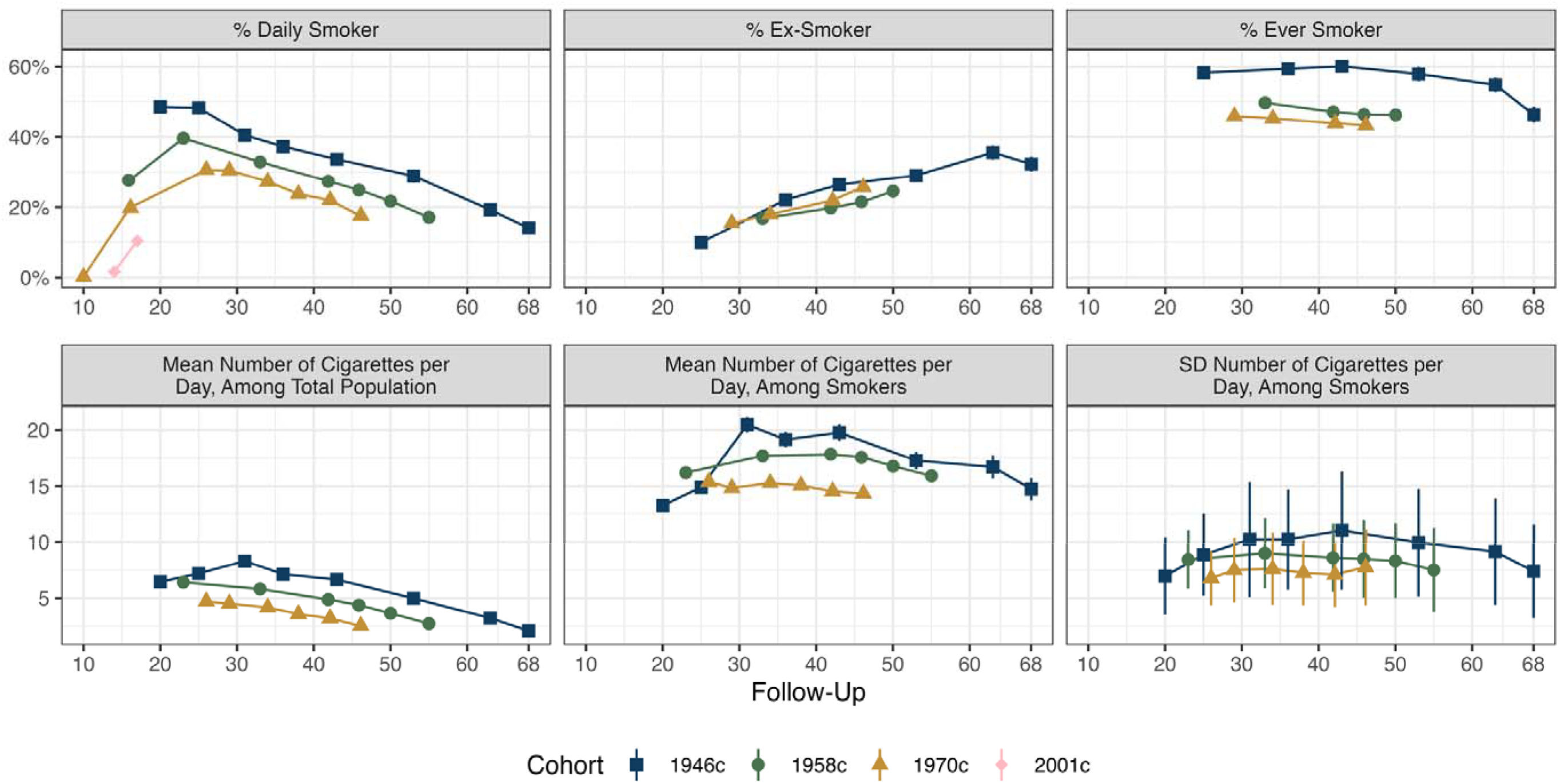
Cohort members’ cigarette smoking, by cohort and age. Calculated using multiply imputed data (m = 40) and accounting for complex survey design.

The average number of cigarettes smoked daily across the total population also declined from early adulthood (bottom left panel, Figure 1). This largely reflected declining prevalence of daily smoking: the number of cigarettes smoked daily *by daily smokers* exhibited weaker change as participants aged (bottom middle panel, Figure 1). Nevertheless, there was evidence that, among *sustained smokers* (i.e. individuals who reported daily smoking in a contiguous run of sweeps), the average number of cigarettes smoked each day declined over time (Supplementary Figure S1).

The average number of cigarettes smoked among smokers was successively lower between cohorts: at age 42/43y, 19.8 (19.0, 20.5) in the 1946c, 17.8 (17.5, 18.1) in the 1958c, and 14.5 (14.2, 14.8) in the 1970c (bottom middle panel, Figure 1). There was also evidence of greater variation in earlier cohorts in the number of cigarettes smoked daily by smokers, though confidence intervals overlapped (bottom right panel, Figure 1).

### Sex-Specific Trends in Cohort Members’ Cigarette Smoking

Men were generally more likely to be daily smokers than women, though these differences varied across time (top panels, Figure 2; see also Supplementary Figures S2-S3). Differences were particularly pronounced in early adulthood in the 1946c: at age 20y, the prevalence of daily smoking was 40.9% (38.4%, 43.5%) among females and 55.6% (53.1%, 58.1%) among males. Differences between sexes later in adulthood in the 1946c and in the latter three cohorts were much smaller. At age 42/43y, the difference in the prevalence of smoking in males compared with females was 3.0 percentage points (pp; 95% CI = -0.7 pp, 6.7 pp) in the 1946c and 0.9 pp (-0.7 pp, 2.6 pp) in the 1958c and 3.7 pp (2.2 pp, 5.2 pp) in the 1970c. The pattern of declining prevalence of smoking after age 30y was observed in both sexes in each cohort (Figure 2; see also Supplementary Figures S1 & S3).

**Figure 2:**
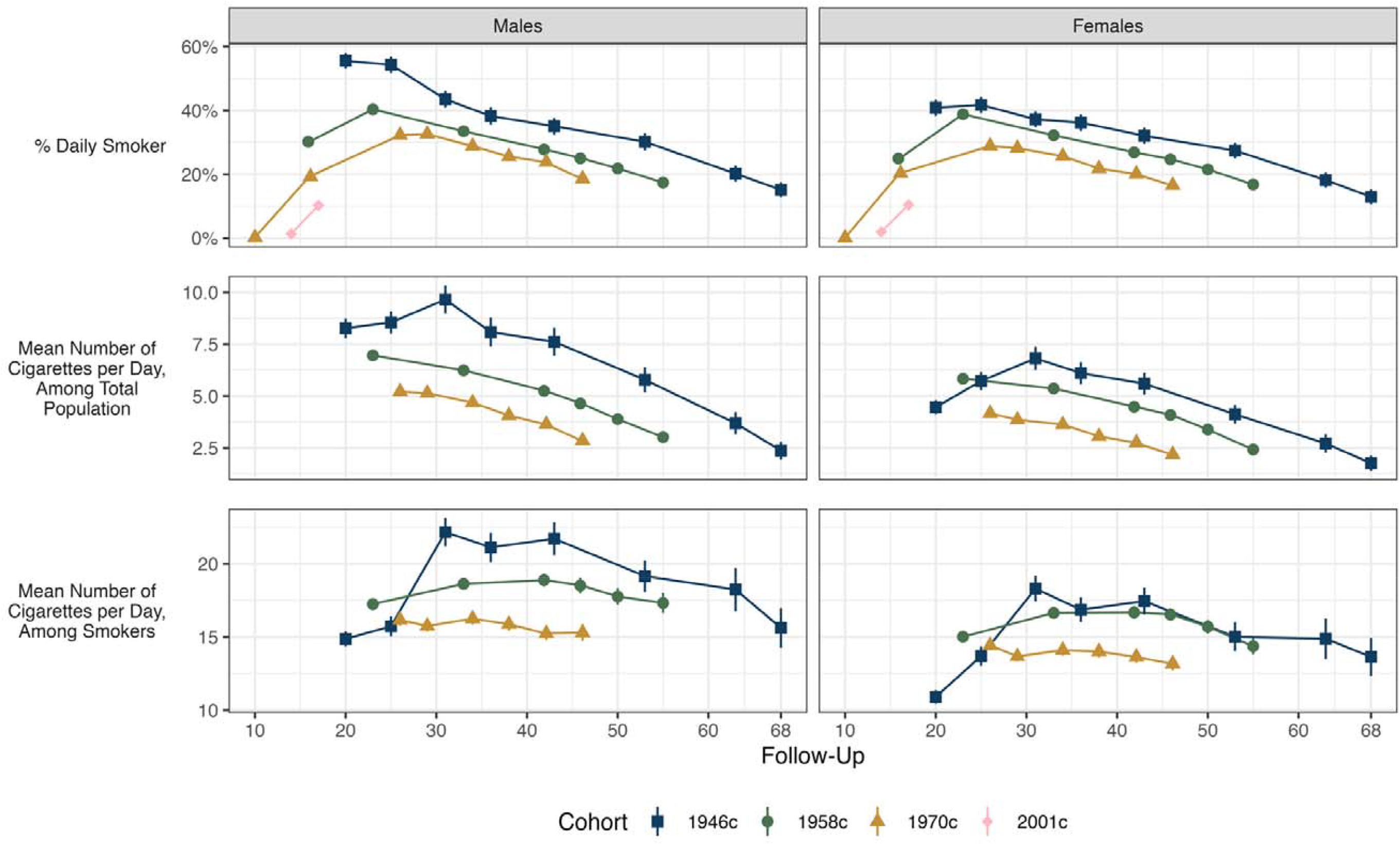
Cohort members’ cigarette smoking, by cohort, sweep and sex. Calculated using multiply imputed data (m = 40) and accounting for complex survey design

The average number of cigarettes smoked daily was higher in men than women in each cohort, both measured across the total population (smokers and non-smokers) and among daily smokers only (middle and bottom panels, Figure 2; see also Supplementary Figures S4-S5). The difference in smoking intensity between male and female smokers was broadly stable across the life course but somewhat small in later cohorts. Among daily smokers at age 42/43y in the 1946c, average number of cigarettes smoked per day by male smokers was 4.3 cigarettes higher (2.8, 5.7) than among female smokers. Corresponding figures were 2.2 (1.6, 2.8) in the 1958c and 1.6 (1.0, 2.2) in the 1970c.

### Parental Cigarette Smoking

Parents were born approximately 30 years before participants on average in each cohort. Maternal smoking during pregnancy was lower in the 2001c than 1958c or 1970c, but rates were higher in the 1970c than 1958c (top panels, Figure 3). 42.3% (41.5%, 43.1%) of participants’ mothers smoked in month 9 of pregnancy in the 1970c, while 33.5% (32.8%, 34.2%) of participants’ mothers smoked in month 4 of pregnancy in the 1958c.

**Figure 3:**
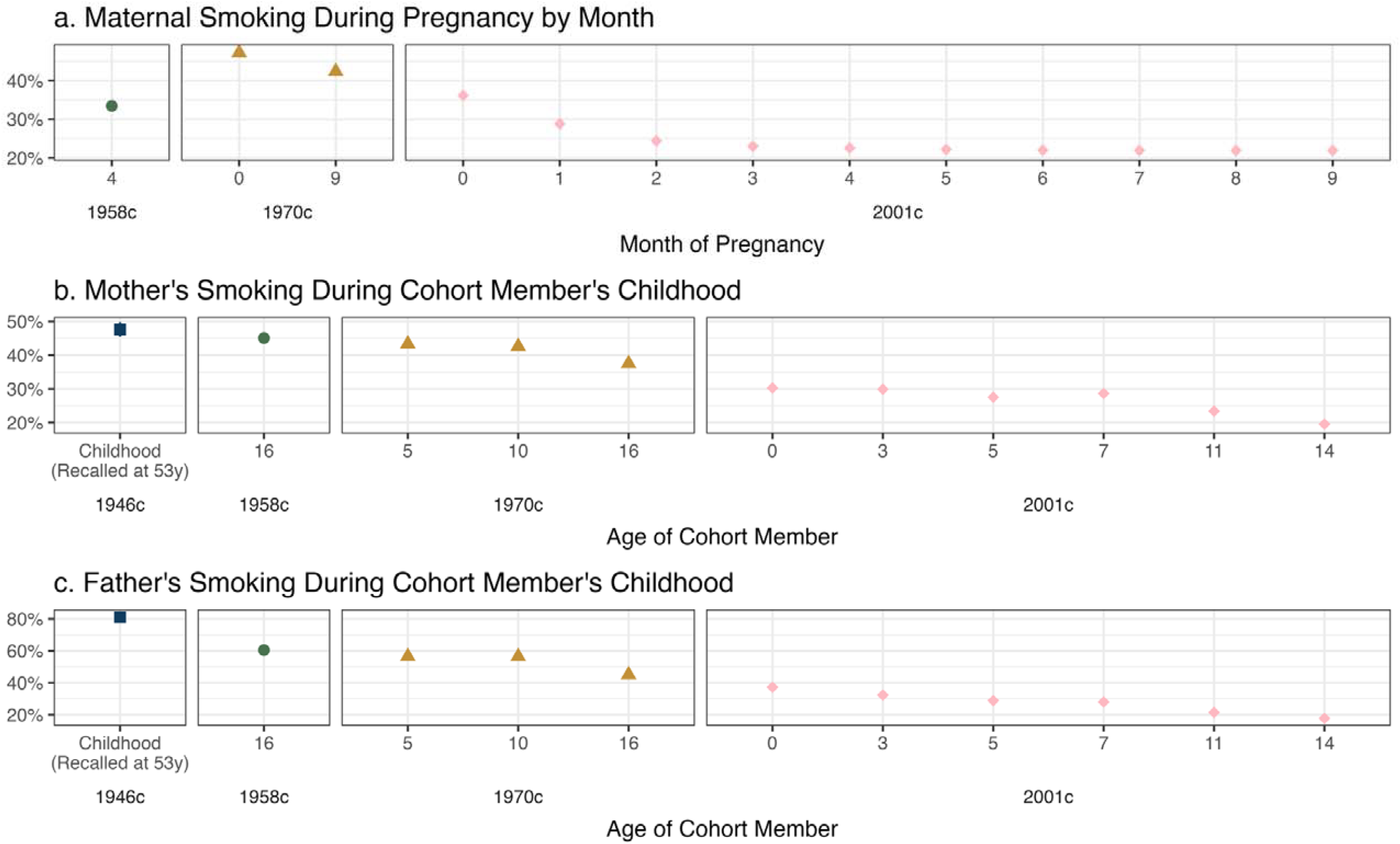
Cohort members’ parents cigarette smoking, by cohort, sweep and sex. Calculated using multiply imputed data (m = 40) and accounting for complex survey design. Note different scales are used for the y-axes.

The prevalence of parental (daily) smoking during participants’ childhood was highest in the 1946c and lowest in the 2001c: 80.5% (78.9%, 82.2%) of fathers smoked in the 1946c, compared with 24.2% (22.8%, 25.5%) at age 14y in the 2001c (middle and bottom panels, Figure 3). Differences between 1958c and 1970c were smaller, especially for mother’s smoking. When participants were age 16y, 45.2% (44.3%, 46.1%) of mother’s smoked in the 1958c and 38.1% (37.2%, 39.0%) in the 1970c.

### Sensitivity Analyses

Results were qualitatively similar when using IPW instead of MI to account for attrition (Supplementary Figures S6). However, in each cohort, smokers had a higher rate of attrition than non-smokers, including at early ages before smoking-related deaths may be expected to occur (Supplementary Figure S7). Though we included smoking observations from earlier and later sweeps in imputation models, data could still be missing not at random (MNAR). Nevertheless, qualitatively similar results were obtained (albeit weaker in size) when adopting the ‘Russell Standard’ and assuming continual smoking for smokers who dropped out – there was lower prevalence of smoking in more recent cohorts and declining prevalence of smoking after early adulthood (Supplementary Figure S8).

## Discussion

We present new harmonised data for smoking behaviour across four British Birth Cohort Studies born 1946-2001. We used these resources to investigate long-run changes in smoking by age and cohort. In each cohort with adulthood data, daily smoking peaked in early adulthood (∼age 25y) and declined thereafter (up to 69y). At a given age, fewer people in more recent cohorts smoked, and those who did, smoked less per day. The prevalence of smoking among participants’ parents was also lower in more recent cohorts. Differences were particularly large among participants’ fathers – for instance, prevalence of smoking declined 55% between the 1946c and 2001c. We also observed sizable sex differences: males smoked more than females and the gender gap decreased between cohorts.

These results are consistent with previous analyses of UK data over shorter timespans (e.g., 1972-2019) that used pseudo-cohorts or retrospective measurement of cigarette smoking (Christopoulou, 2015; Davy, 2006; Kemm, 2001; Opazo Breton et al., 2022). Our results extend these by identifying smaller changes on the intensive margin (i.e., *how* much to smoke), as well as on the extensive margin (i.e., whether to smoke at all); among sustained smokers, average number of cigarettes smoked per day declined as individuals aged, though changes were weaker than changes in the total number of smokers.

As in other studies documenting smoking trends, multiple explanations are possible. For example, declines could reflect changes in beliefs about the health risks of smoking or be largely the consequence of policies designed to reduce tobacco use (e.g., the imposition of taxes). However, we observed largely linear declines in smoking across adulthood, suggesting no single event led to such declines. We also cannot determine whether reductions in cigarette smoking translated into lower ingestion of tar or nicotine – smokers could have responded to higher prices by smoking each cigarette more deeply (Adda & Cornaglia, 2006).

This analysis only scratches the surface of questions that can be asked with these cohort data, including questions on individual smoking trajectories and cross-cohort differences. Rich socioeconomic data (e.g., social class, education, income) enable investigation of changing inequalities in smoking between cohorts and over the life course; detailed health and function data could be used for investigating cohort effects in smoking’s association with morbidity and mortality outcomes; and genetic data (Shireby et al., 2024) could be used to investigate changes in the genetic contribution to smoking at different stages of the tobacco epidemic (Boardman et al., 2010) or before and after specific policy changes, for instance as a test of the ‘hardening hypothesis’ (Beard et al., 2019). Future sweeps (Centre for Longitudinal Studies, 2024b, 2024a, 2024c) may also be useful for investigating changes in vaping, which has become more widespread in recent years (Office for Health Improvement and Disparities, 2022).

### Strengths and Limitations

Strengths include the longitudinal, prospective measurement of cigarette smoking from a nationally representative sample of participants and their parents. Such data mitigate recall biases and allow tracking of smoking behaviour across life. While parental smoking data was collected retrospectively from participants in the 1946c, estimates for fathers’ smoking (∼80%) were very similar to a contemporary data source (Peto et al., 2000).

Limitations include – common with all longitudinal studies – attrition. However, the rich birth cohort data enabled us to draw on early life predictors of attrition and use MI and IPW. Smokers were more likely to drop-out in earlier sweeps. Nevertheless, our results were similar to those identified in cross-sectional data (Opazo Breton et al., 2022) and were qualitatively similar when adopting the very conservative Russell Standard assumption. While the wording of questionnaire items was broadly semantically similar, exact wording often differed, both between cohorts and within a cohort across time, which could have influenced responses. However, our results matched our expectations and were consistent with prior work using other data sources. Finally, to increase comparability between cohorts, we only examined those of white ethnicity. Given the ethnic diversity of the UK has increased over time, our cross-cohort estimates do not capture differences resulting from this change in the composition of the population.

## List of Abbreviations

1946c: National Survey of Health and Development
1958c: National Child Development Study
1970c: 1970 British Cohort Study
2001c: Millennium Cohort Study
95% CI: 95% Confidence Interval
IPW: Inverse Probability Weights
MI: Multiple Imputation
PP: Percentage Points

## Acknowledgements

We would like to thank members of UCL Tobacco & Alcohol Research Group for their detailed comments on a draft version of this manuscript.

## Author contributions

All authors contributed to the analysis plan. LW performed the coding, analysis and data harmonisation and wrote the first draft of the manuscript. All authors provided critical revisions.

## Data availability

The code used to run the analysis is available at https://osf.io/54w6q/. Data from the National Child Development Study, British Cohort Study and Millennium Cohort Study are available through the UK Data Service (https://ukdataservice.ac.uk/). Data from the MRC National Survey of Health and Development are available via application on UCL Skylark (https://skylark.ucl.ac.uk/).

