## Supplementary Information for "Cigarette Smoking Across Life from 1946 to 2018: Harmonisation of Four British Birth Cohort Studies"

### Items on Smoking Behaviour in the Cohort Studies

In this section, we describe the measures on smoking behaviour available in the British Birth Cohort Studies. We also describe measures available in Next Steps (formerly known as the Longitudinal Study of Young People in England; LSYPE; Calderwood & Sanchez, 2016), a representative cohort of English schoolchildren who have been followed since age 13/14y in 2003/04, when they were in Year 9 of secondary school (born 1989/90). Next Steps may be used as an additional data resource for cross-cohort research.

The studies contain a lot of information on smoking. Here, we focus on items on smoking behaviour of cohort members and their parents, partners, cohabitees, and friends. We do not discuss items on recent smoking asked before physical exams (i.e., have you smoked in the last hour?), cigarette manufacturer (asked in some sweeps of the NSHD and BCS70), expenditure on tobacco products, or on general attitudes to smoking and smokers, except for beliefs about the health risks attendant with smoking (questions asked in the BCS70).

Response categories and questionnaire items as mentioned below are conceptual summaries and generally not written verbatim. We separately provide a database of the smoking items, both as an Excel spreadsheet and a searchable online Shiny app (Wright, 2024a, 2024b). This database contains much more information, including exact question wording and response categories, survey modes used, the ‘universe’ of participants particular items were given to, relevant variable names, and the names of these raw dataset files variables are contained in. Data from the NCDS (1958c), BCS70 (1970c), MCS (2001c) and Next Steps are available from the UK Data Service (University College London, 2023a, 2023b, 2023c, 2023d), while data from the 1946c are available by application via the UCL Skylark website (MRC Unit for Lifelong Health & Ageing at UCL, 2024b).

#### National Survey of Health and Development (NSHD, 1946c)

The NSHD commenced before the negative health effects of smoking were well-established (Wadsworth et al., 2006). Cohort members (CMs) turned sixteen years old in the week the Royal College of Physicians (1962) released their landmark report, *Smoking and Health*, which detailed evidence on the link between smoking and lung cancer, among other illnesses (James, 2024). As a result, data on smoking behaviour has only been captured from age 20y onwards in the NSHD, including data on parental smoking, which was only collected retrospectively from CMs at age 53y. There is no data on smoking during pregnancy, either for the CM’s mothers or for female CMs themselves.

Items on CMs’ smoking behaviour were included at the age 20y, 25y, 31y, 36y, 43y, 53y, 63y, 68y, and 69y sweeps. At age 20y, CMs were asked how frequently they currently smoked cigarettes (regularly, less than once per day, or no), what they smoked (cigarettes, roll-ups, cigars and pipes), whether they inhaled, and the amount smoked (cigarettes per day, ounces of tobacco a week, or cigars per week). Smokers were also asked the number of cigarettes they smoked per day a year ago, when they started smoking, and the maximum they had smoked a day for as long as a year. Non-smokers were asked if they had ever smoked cigarettes (regularly, less than once per day, no), the maximum amount smoked for as long as a year, whether they inhaled, when they began smoking, and how old they were when they stopped and why. Similar questions were asked at 25y, though ex-smokers were asked specifically how much they were smoking when they gave up, and regular smoking was unambiguously defined (1+ cigarettes per day over previous month for current smokers, and 1+ cigarettes per day for as long as a year for ex-smokers). CMs were also not asked about smoking one year ago and non-smokers were not asked why they stopped.

At 31y, CMs were asked whether they were regular, occasional, ex-, or non-smokers (terms not explicitly defined) of cigarettes, cigars or pipes, how far they take the smoke in, when they last gave up cigarette smoking for a month or more, and the usual number of cigarettes smoke over the last year or, for ex-smokers, the number smoked per day before giving up. Items on smoking at ages 20y, 25y and 31y were all obtained via self-complete postal questionnaire.

At 36y, 43y, and 53y, smoking behaviour was elicited via face-to-face interview. At 36y and 53y, participants were asked whether they currently smoked cigarettes, cigars, or a pipe, whether they inhale, and for the amount smoked of each (cigarettes per day, cigars per week, ounces of pipe tobacco per week). At age 36y, non-smokers were asked whether they had ever smoked daily for as long as a year, why they quit and when. Due to routing, it is not possible to determine whether current irregular smokers had ever regularly smoked. This is also the case at age 20y, 63y and 68y. At ages 43y and 53y, the question on ever smoking was instead asked of all participants. At age 43y, CMs were additionally asked whether they had tried to quit and when. These questions were not asked at age 53y, but CMs were given two items on whether anyone they lived with smoked and the average number of cigarettes household members smoked at home per day at this sweep. At age 53y, CMs were also asked a retrospective item on parental smoking during childhood (“Did either of your parents smoke cigarettes, cigars or pipes when you lived with them as a child? Mother, father, neither, both”).

At ages 63y and 68y, the survey reverted to self-complete postal questionnaire. At 63y, CMs were asked (in separate questions) whether they smoke cigarettes, cigars, or a pipe at all nowadays, the amount smoked (e.g., cigarettes per day), age at starting, and, for non-smokers, whether they had ever smoked regularly and when they quit. At 68y, four items were included on current smoking at all, cigarettes per day, whether the participant had ever smoked (current non-smokers only) and, if so, when they quit. Finally, at 69y, which was conducted via face-to-face interview, participants were only asked whether they smoked at all nowadays.

During the COVID-19 pandemic, cohort members were also invited to complete three surveys, each of which contained information on smoking behaviour (MRC Unit for Lifelong Health & Ageing at UCL, 2024a; University College London, 2024). The same survey was given to the other birth cohort studies and Next Steps. Consequently, we describe the smoking-related items in a separate section (*COVID-19 Studies*) below.

Data from the NSHD are obtained via application. The NSHD operate a bespoke dataset model, releasing only variables that are requested and related to an applicant’s project. We have created a ‘basket’ of smoking variables (Basket ID = ljwright90ZZvelweu), which can be viewed on UCL Skylark, the NSHD’s data discovery system (MRC Unit for Lifelong Health & Ageing at UCL, 2024b), and used for researcher’s own data applications.

#### The National Child Development Study (NCDS, 1958c)

Data for the NCDS are currently available up to age 55y. Data has been collected on the smoking behaviour of CMs, their parents and their partners at multiple sweeps. At age 0y, the CM’s mother was asked about the number of cigarettes she smoked per day in the 12 months prior to pregnancy and for changes in smoking during pregnancy, including the month of the change and the number smoked per day thereafter. From this, the data managers have created a composite derived variable on smoking after month 4 of the pregnancy.

Data on the amount smoked by the CM’s mother and father per day was collected at age 16y from the responding parent (typically the CM’s mother). Responding parents were additionally asked whether they had argued with the CM about the amount the CM smokes. This sweep also contained a single self-completion item given to CMs on cigarettes smoked per week (none, < 1, 1-9, 10-19, 20-29, …, 50-59, 60+). Items given to the responding parent were instead elicited via face-to-face interview.

At age 23y, CMs were asked whether they had ever smoked, whether they smoked cigarettes at all currently, the amount they smoked per day on average, and for non-smokers, whether they had ever smoked regularly (1+ cigarettes a day for 12+ months) and how old they were when they stopped. As with the NSHD, the routing of this question meant that irregular smokers were not asked whether they had ever smoked regularly. Also at this sweep, female smokers and ex-smokers who had had a baby were asked, for their most recent baby, whether they had smoked prior to pregnancy and what amount, whether and when they had changed the amount smoked during pregnancy, and for the amount smoked after the change. All CMs were also asked whether they currently lived with anyone who smoked, who that person was, and, if their partner smoked, the number of cigarettes they smoked per day. Each of the questions at age 23y were asked via face-to-face interview.

The questions on household members’ smoking were also asked at age 33y. At this sweep, participants were also asked whether they smoked at all nowadays, the amount they usually smoked per day, and whether they had ever smoked regularly. Unlike at age 23y, the latter question was asked of irregular smokers, too. Former smokers were asked how many cigarettes they usually used to smoke and the age at which they quit. The sweep also contained two sections on smoking during pregnancy. The first section was asked of all CMs (male or female) who had conceived a pregnancy. As at 23y, participants were asked whether and the amount they smoked prior to the pregnancy, whether and when they had changed the amount they smoked during pregnancy, and for the amount smoked after the change. Unlike at age 23y, these questions were repeated for each pregnancy. The second section was on smoking during pregnancy was given to natural mothers of a selected CM’s child; for males, this was the CM’s partner, while for females, this was the CM themselves. For the selected child, the natural mother was asked the amount smoked prior to pregnancy and the average amount smoked per day after the third month of pregnancy. Each of the items on smoking at the age 33y sweep were given via face-to-face interview.

Questions on smoking during pregnancy were also asked at age 42y for all pregnancies that had occurred since the age 33y sweep, and again asked of male and female CMs. A more limited set of items was given, however: CMs were asked whether the participant had smoked in the 3 months before pregnancy, in months 1-5 and months 6-9, and whether they had smoked more, less or the same during pregnancy as before. At age 42y, participants were also asked for their current smoking frequency (never, used to but no more, occasionally, every day) and amount (cigarettes per day). Non-regular smokers were asked if they had ever smoked regularly (1+ cigarettes for 12+ months), how old they were when they stopped. The questions from the ages 23y and 33y sweeps on household member and partner smoking habits were again asked.

The same questions from age 42y, except for those on smoking during pregnancy and household and partner smoking, were asked at ages 46y and 50y. At 50y, current and former smokers were additionally asked at what age they had begun to smoke. At age 55y only two items on smoking behaviour were given – on currently smoking frequency (every day, occasional, used to, never) and amount (cigarettes a day), respectively. In latter sweeps, the BCS70 and NCDS have been co-developed with many of the same survey items given to both cohorts, facilitating harmonization (Joint Centre For Longitudinal Research, 2001).

During the COVID-19 pandemic, cohort members were also invited to complete three surveys, each of which contained information on smoking behaviour (University College London, 2024). The same survey was given to the other birth cohort studies and Next Steps. Consequently, we describe the smoking-related items in a separate section (*COVID-19 Studies*) below.

The NCDS also includes a sub-study performed for the Tobacco Research Council when participants were age 20y (National Children’s Bureau, 2023). We do not discuss the items available in this study further as it was only carried out on ~ 800 participants (<5% of the initial sample) and the data deposited on the UKDS have not been cleaned.

#### 1970 British Cohort Study (BCS70, 1970c)

Data for the BCS70 are currently available up to age 46y. Items on smoking have been included in every sweep. Data on smoking have been collected on the smoking behaviour of CMs, their parents, friends and partners.

At age 0y, CM’s mothers were asked whether they currently smoked, or had ever smoked, how much they smoke(d) and, if a non-smoker, when they had stopped. They were also asked whether they smoked during pregnancy. From these items, the data owners provide a composite derived variable on smoking during pregnancy, including cigarettes smoked. The 1970c also contains a substudy when participants were 22 months old, which included items on maternal smoking, including during pregnancy. However, data is only available for ~ 2,500 cohort members (< 15% of the total sample), so we do not discuss these items further here (see database for more information).

At age 5y, CM’s mothers were asked (separately) whether they or the CM’s father were regular smokers of cigarettes (1+ per day, on average), a pipe, or cigars, and how many cigarettes they smoked per day on average. They were also asked for how long since the CM’s birth each parent had smoked. CM’s mothers were again asked at the age 10y sweep whether they or the CM’s father currently smokes and for the number of cigarettes smoked per day. The mother was also asked how long each parent had smoked, and if not smoking currently, whether they had smoked in the past ten years, and for the amount smoked before giving up and the time since quitting. Additionally, the CM’s mother was asked whether and how many other members of the household currently smoked. Items given to the mother at ages 0y, 5y and 10y were via face-to-face interview.

The age 10y sweep also contained four items given to CMs. Unlike the items given to the mother, CM’s responses were made by self-completion. CMs were asked whether they had tried a one or more than one cigarette, and for their current smoking amount and frequency (never tried, tried once, tried twice, < 1 per week, ~ 1 per week, 2-5 per week, ~ 1 a day, > 1 a day). CMs were also asked whether they believed smoking harm’s people’s health. A more extensive set of items was given to CMs at age 16y, some of it repetitive, and including items on the CM’s attitudes and beliefs about smoking and *smokers*. For instance, CMs were asked for the level of agreement with seventeen items on smoking, including seven on its health risks (“Smoking is only bad for you if you do it for years”, “Breathing other people’s smoke harms non-smokers”, “Most people who get lung cancer have smoked regularly”, “Smoking is only bad for you if you smoke a lot”, “If a woman smokes when she is pregnant it may harm her baby”, “Smokers live as long as non smokers”, “Some cigarettes are not dangerous”).

Regarding smoking behaviour, at age 16y, CMs were asked how many cigarettes they had smoked since yesterday and last week. They were also asked their smoking status in two separate items. The first allowed responses for never smoked, tried once or twice, smoked sometimes but not now, current smoke and would like to give up, and do not want to give up smoking. The second allowed responses for never smoked a cigarette, used to smoke but not for the past 3+ months, smoke less than once a week, and smoke one cigarette per week. CMs were also asked for the amount smoked per week (non-smoker, 1, 2-5, 6-10, 11-20, 21-40, 41-70, 70-100, > 100), their age at first trying a cigarette, how their smoking habit had changed over the past year, and whether they expected to be smoking in twelve months, in addition to a set questions related to specific behaviours such as amount inhaled (see attached database for all items at this sweep). CMs were further asked how often their mother, father, siblings, best friend, and partner smoke d(not at all, sometimes, often) and asked how many of their friends smoked. Note, there was a national teacher strike during the age 16y sweep, which led to unexpectedly low response rates at this sweep (Elliott & Shepherd, 2006).

CM’s mothers also answered items on smoking behaviour at the age 16y sweep. Specifically, they were asked for themselves, their partner, and the CM, whether they smoked daily or did in the past (and what), how much they currently smoked, when they started, and for non-smokers, when they stopped and how much they smoked before they quit. All items at the age 16y sweep were obtained via self-completion.

At age 26y, CMs were asked for their smoking frequency (never, used to but not now, occasionally, every day) and amount smoked (cigarettes and cigars per day, obtained separately). Data from the 26y was also obtained via self-completion. From age 29y to 46y, face-to-face interview or telephone survey was used, with an exception for one item. At ages 29y and 34y, CMs were again asked for smoking frequency and amount smoked, though the number of cigars was not elicited. Whether the CM had ever smoked regularly (1+ cigarettes per day for 12+ months) was also obtained, as well as their age at quitting. CMs were additionally asked whether any household members smoked, who, and for the amount their partner smoked per day.

At ages 29y and 34y, CMs who had conceived a child were also asked whether they had smoked 3 months prior to pregnancy, in months 1-5 and months 6-9, and, if smoking during pregnancy, whether this was less or more than beforehand. These questions were repeated for each pregnancy but were only asked of females at age 34y. The age 34y sweep also contained a self-complete item on smoking frequency given to a selected CM’s child aged 10-16 years old (never, once or twice, used to but not now, occasionally, 1+ times per week).

The questions on CM smoking frequency, smoking amount, ever smoking regularly and (for ex-smokers) age at quitting were again asked at ages 38y, 42y and 46y. Ages 42y and 46y also contained a question on age the CM starting smoking regularly, which was given to current and ex- daily smokers. Finally, the age 46y sweep contained items on e-cigarette use (never, former, occasional, every day) and on partner’s cigarette smoking frequency (never, former, occasional, every day). As mentioned in the previous section, in later years, the BCS70 and NCDS have been co-developed with many of the same survey items given to both cohorts, facilitating harmonization (Joint Centre For Longitudinal Research, 2001).

During the COVID-19 pandemic, cohort members were also invited to complete three surveys, each of which contained information on smoking behaviour (University College London, 2024). The same survey was given to the other birth cohort studies and Next Steps. Consequently, we describe the smoking-related items in a separate section (*COVID-19 Studies*) below.

#### Millennium Cohort Study (MCS, 2001c)

Seven sweeps of data are currently available for the MCS: at 9 months (9m), 3y, 5y, 7y, 11y, 14y, and 17y, respectively. Items on smoking have been included at each sweep. The MCS has collected information on smoking from CMs, their parents, and selected older siblings. At the first sweep (carried out when cohort members were 9 months old), biological parents (mothers and fathers) were asked for their smoking amount just before the CM’s mother became pregnant (cigarettes per day), whether and when the parent changed the amount they smoked during the pregnancy, and for the amount they smoked after this change. This information was obtained via face-to-face interview.

Between 9m and 14y, parents and carers were asked for the tobacco products they used (cigarettes, roll-ups, cigars, pipe, other), the current number of cigarettes or roll-ups they smoked per day, and whether they had ever smoked cigarettes regularly (once per day for 12+ months). At the 9m sweep, parents were additionally asked for their frequency of using other tobacco products and whether they had smoked in the previous two years, the latter of which was used as a filter for questions on pregnancy during smoking. Between 3y to 11y, the item on ever smoking regularly was limited to parents who had not previously answered this question, and the question was not included at all at the age 14y sweep. Note, a potential issue with the filters is that current irregular smokers were not asked the question on ever being a regular smoker. At the 9m to 11y sweeps, the main parent was additionally asked whether anyone smokes in the same room as the cohort member.

The 3y and 5y sweeps additionally contained questionnaires given to selected older siblings. Siblings aged 10-15y were asked (via self-report) whether they ever smoke cigarettes and for the number of cigarettes they had smoked in the past seven days. Main parents were additionally asked, for selected siblings aged 8-15, whether they thought the sibling had had a problem with smoking in the past 12 months.

Questions on cohort member’s smoking were given in in 11y to 17y sweep, each collected by self-report. At the age 11y sweep, CM’s were asked how many of their friends smoked and whether they had ever tried a cigarette (including a single puff). At the 14y sweep, CM’s were again asked many of their friends smoked. They were also asked whether for their current smoking status (never, tried once, used to sometimes, less than once a week, 1-6 per week, 7+ per week), their age at first trying a cigarette, and their current use of e-cigarettes (never tried, used but not now, occasional use, every day). Similar questions on CM’s cigarette smoking and e-cigarette use were also asked in Sweep 7 (17y), except the response categories for e-cigarette use changed and reflected the response categories for cigarette smoking.

During the COVID-19 pandemic, cohort members and their parents were also invited to complete three surveys, each of which contained information on smoking behaviour (University College London, 2024). The same survey was given to the other birth cohort studies and Next Steps. Consequently, we describe the smoking-related items in a separate section (*COVID-19 Studies*) below.

#### Next Steps (1989/1990)

Data for Next Steps are currently available for nine sweeps from age 13/14y to 32y. Items on CM’s smoking were included in Sweeps 1-3 (ages 13/14y, 15/16y, and 17/18y) and at Sweeps 8 & 9 (25y and 32y). All items were collected in self-completion modules. No information on parental smoking has been collected. Sweeps 1-3 each contained two items, which were repeated at these sweeps: one on smoking at all (“Do you ever smoke cigarettes at all? Yes/No”), and the second mixing smoking amount and frequency (never, ever tried once, used to smoke sometimes, less than once a week, 1-6 cigarettes per week, 7+ cigarettes per week). Unfortunately, these items have issues. The first item uses the present tense and captures current behaviour, but only those who respond “Yes” are shown the second item which attempts to collect information on experience (ever smoked) and current frequency and smoking amounts simultaneously. It is thus unclear from these items who has never smoked (it is possible to answer no to the first question but have tried cigarettes) and who is, e.g., a daily smoker (though an assumption could be made that a person who smokes more than six cigarettes a week, smokes at least one per day).

At ages 25y and 32y, CMs were asked for information on current frequency (never, no longer, occasionally, every day) and amount smoked (cigarettes per day), whether the CM had ever smoked regularly (once per day for at least 12 months), and their ages of starting and stopping smoking regularly. Participants were also asked about frequency of e-cigarette use (never tried, tried but no longer, occasionally, every day), and at age 32y, for the amount of time spent smoking e-cigarettes each day (< 5 minutes; 5-30 minutes; 30 minutes-1 hour; 1-2 hours; > 2 hours). Note, the response categories for the question on smoking frequency are vague in that they do not clarify whether a participant has never *tried* cigarettes or whether they have never smoked regularly.

During the COVID-19 pandemic, cohort members were also invited to complete three surveys, each of which contained information on smoking behaviour (University College London, 2024). The same survey was given to the four British Birth Cohort Studies. Consequently, we describe the smoking-related items in the next section.

#### COVID-19 Studies

The UK entered lockdown in March 2020 (Hale et al., 2020). The Centre for Longitudinal Studies (CLS), which manages the NCDS, BCS70, MCS, and Next Steps, and the MRC Unit for Lifelong Health & Ageing Unit (MRC LHA), which manages the NSHD, collaborated to run three waves of a COVID-19 Survey, with the same questionnaire items given to each cohort. Wave 1 ran from May-June 2020 and, given lockdown restrictions, was carried out via web questionnaire and only offered to cohort members for whom an email address was available; though as restrictions eased, the NSHD were also offered a postal questionnaire, which ran from June-July 2020. Wave 2 was carried from September-October 2020, again via web questionnaire, with 1946c cohort members sent a postal questionnaire between November-December 2020, if they had not responded. The Wave 2 survey was offered to all participants, including those without an email address. Wave 3 ran from February-March 2021. All cohorts were initially invited to participated via web. Non-respondents for CLS’ cohorts were invited to participate by telephone, while a postal questionnaire was instead used for non-respondents in the NSHD (May-July 2021). For the MCS, both cohort members and parents were invited to take part. Only parents who had responded to the mainstage 17y sweep were invited for Wave 1, but all parents were invited thereafter.

Each wave contained a similar set of items on smoking cigarettes and using electronic cigarettes or vaping devices. Regarding cigarettes smoking, participants were asked how frequently they smoked cigarettes currently (never, no longer, occasionally, every day), for the number of cigarettes smoked per day in the month before the COVID-19 outbreak in March 2020, and the number smoked since the start of the pandemic (Wave 1) or in the four weeks prior to completing the survey (Waves 2 and 3). Regarding electronic cigarettes, participants were asked for their frequency of current use, and whether and how this had changed (more, less, the same) since the start of the pandemic (Wave 1) or over the past four weeks (Waves 2 and 3).

### Notes on Harmonisation

There are several differences between cohorts that add complexity to harmonising the smoking data. This includes differences in sampling, data collection, and the survey items given to participants. We outline the important differences here.

#### Sampling

The NSHD is a stratified sample of singletons born in mainland Britian (England, Scotland, and Wales) in a single week of March 1946; individuals from advantaged socioeconomic backgrounds were oversampled. The sample is static – boost samples have not been recruited at later dates. The NCDS and BCS70 are birth cohorts of individuals born in a single week of March 1958 and April 1970, respectively. Participants in the NCDS were born in Great Britain (including offshore islands), while those in the BCS70 were born across the UK, though participants from Northern Ireland were not followed after the first sweep. Both the NCDS and BCS70 are not fully static samples: immigrants to the UK born in the same weeks as the original samples were recruited during childhood using school enrollment information.

Participants in the MCS were recruited from across the UK using a stratified sampling design (oversamples of ethnic minorities and individuals from Scotland, Wales or Northern Ireland, or from disadvantaged backgrounds). Approximately 700 families were recruited at the second sweep (3y) as they were missed by the initial sampling frame. All participants have been eligible for follow-up. Participants in Next Steps were recruited from schools using a two-stage stratified design (ethnic minorities and individuals from disadvantaged backgrounds oversampled). A boost sample of 352 Black ethnicity individuals was added in Sweep 4 (age 16/17y). Individuals have been followed across the UK thereafter.

The cohorts thus differ on the countries individuals have been sampled from. Further, the ethnic make-up of the samples has changed considerably over time. Participants in the 1946c can be presumed to be White, while a sizeable proportion of MCS participants are from ethnic minority backgrounds. Depending on the purpose of the investigation, this can influence cross-cohort trends. In this study, we focus on singletons of White ethnicity born in mainland Britain (England, Scotland or Wales) to ensure comparability across the cohorts used. We also use survey weights to restore representativeness of the samples.

#### Data Collection

The cohorts also differ in the ages that smoking behaviour has been measured. As smoking behaviours change within individuals over the life course (Christopoulou, 2015), comparing the cohorts at different ages may bias estimates of cross-cohort differences. Figure S9 shows the ages in each of the cohorts at which CM smoking behaviour (top panel) and CM parental smoking behaviour (bottom panel) have been collected. There are few ages at which cohorts overlap. Importantly, there is overlap in the NSHD, NCDS and BCS70 at age 42/43y – this age has previously been used for cross-cohort examinations and harmonization efforts (e.g., McElroy et al., 2021). We also exploit this here by focusing some analyses on items collected at these sweeps.

An additional difference in the data collection procedures is the survey modes used. This has also varied within studies over time. For instance, many of the earlier smoking items have been collected using self-complete postal questionnaires, while more recent sweeps have used computer assisted face-to-face, web, or telephone interviews. (It is worth noting that, for postal questionnaires, routing between questions cannot be enforced – e.g., answering questions on age at quitting smoking only after revealing oneself as a former smoker. This leaders to a small number of discrepancies between variables, decisions about which need to be taken.) A large literature has investigated mode effects, showing that responses can differ systematically across survey modes (e.g., Jäckle et al., 2010; Maslovskaya et al., 2023). Importantly, given smoking is often seen as an undesirable behaviour (East et al., 2019), it is possible that reporting rates could be lower in non-anonymous modes, such as face-to-face interviews. However, it should be noted, there was no evidence of differences in responses across mode the age 55y sweep of the NCDS, which embedded a mode effects experiment with people randomized to respond via (non-anonymous) telephone or (anonymous) web survey (Goodman et al., 2022).

#### Survey Items

More obviously, the survey items differ between cohorts and within a cohort, across time. This includes the tobacco products that are enquired about – in earlier sweeps of the NSHD, cohort members are asked about pipe and cigar smoking, but this is largely absent elsewhere. Each study contains items on smoking frequency (e.g., occasional or daily use) and smoking amounts (e.g., cigarettes smoked per day), but these differ on the precise wording of the questions, including the time frames adopted (weekly or daily use) and on how responses are categorized (e.g., some sweeps ask for specific number of cigarettes smoked, others given ranges in categories). Additionally, some survey sweeps use precise language to define behaviours (e.g., regular smoking as smoking at least one cigarette per day for twelve or more months). Others use imprecise language, and when combined with specific question routing, can lead to non-exhaustive categorisations being used – for instance, occasional smokers not being asked whether they have ever smoked regularly. This is also evidence in questions like ‘Do you smoke cigarettes?’ and ‘How old were you when you began smoking cigarettes?’ which a person may interpret as meaning regular use but could also mean occasional use or recently trying. Each of these makes harmonization fraught.

### Smoking Measures used in the Present Study

#### Cohort Member Current Daily Smoking

We defined current daily smoking as smoking one or more cigarettes per day at the time of interview, focusing on mainstage survey sweeps rather than the COVID-19 surveys. Daily smoking can be identified either through questions on smoking status (with ‘smoking every day’ or similar appearing as an option) or on smoking intensity (e.g., cigarettes smoked per day). In sweeps where questions on both smoking status *and* smoking intensity appeared, we prioritized the former, given the latter may misidentify individuals who smoke less regularly: in some sweeps, the provision of an option to report smoking less than one cigarette per day was not explicit or even not permissible. For instance, at ages 36y and 53y in the NSHD, valid responses were restricted to 1 or more cigarettes.^[1]^ Further, occasional smokers could have plausibly interpreted questions on smoking intensity as enquiring about cigarette consumption on days that they smoked; in the NSHD, at age 20y, almost half of occasional smokers (according to smoking status) reported smoking 1 or more cigarettes a day. Nevertheless, the total number of occasional smokers was relatively small, and not using smoking intensity would have meant losing data from multiple sweeps (ages 23y-33y in the NCDS and ages 36y-68y in the NSHD), so we opted to use data on smoking intensity where no better option was available.

Questions on smoking intensity were sometimes restricted to categorical responses (e.g., 0 cigarettes, 1-5 cigarettes, 5-10 cigarettes, …) or elicited consumption over timeframes longer than a day (e.g., cigarettes per week). We convert longer timeframes to daily amounts (e.g., dividing weekly amounts by seven), which may have led to incorrect classification of individuals who did not smoke consistently. For intensity amounts reported categorically, we took a conservative approach, limiting to categories where every possible response entailed smoking 1+ cigarettes per day, on average (e.g., 5-10 cigarettes per week *would not* count as daily smoking). In sensitivity analyses, we relaxed this assumption and classified as daily smoking any response that could potentially have entailed smoking 1+ cigarettes per day, on average (e.g., 5-10 cigarettes per week *would* count as daily smoking). Results were substantively similar when doing so (results available upon request).

Survey items were based on self-report in all cases, but survey mode differed between cohorts and sweeps, including via non-anonymous (face-to-face or telephone interview) and anonymous (paper or web self-completion) modes. It is possible that lack of anonymity could influence influences responses, particularly where smoking is considered socially unacceptable, for instance by reducing the likelihood a participant admits smoking or increasing the likelihood that they underreport smoking intensity. However (as noted above), there was no evidence of an effect of mode on responses at the age 55y sweep of the NCDS, which embedded a mode effects experiment with people randomized to respond via (non-anonymous) telephone or (anonymous) web survey (Goodman et al., 2022).

Note, unlike for former smoking (see next section), in almost all sweeps, questions on *current* smoking did not specify a timeframe over which the behaviour had to be sustained. It is thus possible for a participant to be classified as current smoker in one sweep but not an ex-smoker in a later sweep, even though they had since quit. It is difficult to gauge this possibility empirically – questions on age started smoking regularly were not always asked and not reporting previous smoking could also reflect recall bias.

We created daily smoking variables for ages 20y, 25y, 31y, 36y, 43y, 53y, 63y, and 68y in the NSHD, ages 16y, 23y, 33y, 42y, 46y, 50y, and 55y in the NCDS, ages 10y, 16y, 26y, 29y, 34y, 38y, 42y, and 46y in the BCS70, and ages 14y and 17y in the MCS. The variables and specific definitions used to define current daily smoking are shown in Table 1below.

Table 1: Variables used to define current daily smoking by cohort-sweep.

| **Cohort** | **Sweep** | **Definition (Case)** | **Variables** | **Notes** |
| --- | --- | --- | --- | --- |
| MCS | 14y | Responds to question on smoking status with ‘6. I usually smoke more than six cigarettes a week’ | FCSMOK00 |  |
| MCS | 17y | Responds to question on smoking status with ‘6. I usually smoke more than six cigarettes a week’ | GCSMOK00 |  |
| BCS70 | 10y | Responds to question on current smoking with ‘7. Smoke about 1 cigarette a day’ or ‘8. Smoke more than 1 cigarette a day’ | k051, k052, k053 |  |
| BCS70 | 16y | Responds to question on amount smoked per week as ‘5. More than 10 and up to 20’ or higher (gh1). | gh1, gh2 | One lower category is ‘More than 5 and up to 10’, which also includes people smoking an average of 1+ per day |
| BCS70 | 26y | Responds to question on current smoking habit as ‘4. I smoke every day’. | b960632 |  |
| BCS70 | 29y | Responds to question on current smoking status as ‘4. you smoke cigarettes every day?’ | smoking |  |
| BCS70 | 34y | Responds to question on current smoking status as ‘4. you smoke cigarettes every day?’ | b7smokig |  |
| BCS70 | 38y | Responds to question on current smoking status as ‘4. you smoke cigarettes every day?’ | b8smokig |  |
| BCS70 | 42y | Responds to question on current smoking status as ‘4. I smoke cigarettes every day’ | B9SMOKIG |  |
| BCS70 | 46y | Responds to question on current smoking status as ‘4. I smoke cigarettes every day’ | B10SMOKIG |  |
| NCDS | 16y | Responds to question on amount smoked per week as ‘4. 10 to 19 a week’ or higher | n2887 | One lower category is ‘1 to 9 a week’, which also includes people smoking an average of 1+ per day |
| NCDS | 23y | Responds to question on cigarettes usually smoked per day as 1 or more. | n5930, n5931, n5935 |  |
| NCDS | 33y | Responds to question on cigarettes usually smoked per day as 1 or more. | n504262, n504263 |  |
| NCDS | 42y | Responds to question on current smoking status as ‘4. you smoke cigarettes every day?’ | smoking |  |
| NCDS | 46y | Responds to question on current smoking status as ‘4. you smoke cigarettes every day?’ | n7smokig |  |
| NCDS | 50y | Responds to question on current smoking status as ‘4. you smoke cigarettes every day?’ | N8SMOKIG |  |
| NCDS | 55y | Responds to question on current smoking status as ‘1. I smoke cigarettes every day’ | N9SMOKIG |  |
| NSHD | 20y | Responds to question on current smoking status as ‘1. Yes, regularly’ | SMO66 | Also asks questions on pipe and cigars, which we have not included. Question defines occasional smoking explicitly as usually less than 1 cigarette per day. |
| NSHD | 25y | Responds to question on regular smoking as ‘1. Yes’. | SMO71, SMOD71,  SMOW71, SMOR71 | Regular smoking is defined explicitly (1+ cigarettes per day or 1oz rolling tobacco total over the past month). Could potentially get confused with pipe smoking |
| NSHD | 31y | Responds to question on current smoking status as ‘3. a regular smoker of’ | SMOS77 | Regular smoking is not defined implicitly (as at age 20y) or explicitly (as at age 25y). |
| NSHD | 36y | Responds to question on amount smoked per day as 1 or more | SMOS82, SMODS82 | There does not appear to be an option for less than 1 per day, which could mix occasional and regular smokers. |
| NSHD | 43y | Responds to question on amount smoked per day as 1 or more | SMOS89, SMODS89 |  |
| NSHD | 53y | Responds to question on amount smoked per day as 1 or more | SMOS, SMODS | There does not appear to be an option for less than 1 per day, which could mix occasional and regular smokers. |
| NSHD | 63y | Responds to question on amount smoked per day as 1 or more | SMO09, SMOD09 |  |
| NSHD | 68y | Responds to question on amount smoked per day as 1 or more | SMO14x, SMOD14x |  |

#### Cohort Member Current Smoking Intensity

We defined current smoking intensity as cigarettes smoked per day. Question wording typically enquired about ‘usual’ daily consumption (BCS70 and NCDS) or consumption ‘now’ (NSHD), and generally did not specify a period over which the behaviour had to be sustained (age 31y in the NSHD was an exception).

Where the timeframe was longer than a day, or where different days were enquired about (e.g., weekday and weekend), we converted responses into an average daily amount (e.g., dividing weekly amount by seven). Early sweeps of the NSHD also contained items on ounces (oz) of rolling tobacco consumed per week. We converted this to an equivalent number of cigarettes assuming 1oz of rolling tobacco was made 40 cigarettes (1oz per week = 5.8 cigarettes per day). Given the lack of granularity, we disregarded items where smoking intensity was categorized (e.g., 1-5, 6-10, …, 60+ cigarettes per day), except for the NSHD items on rolling tobacco consumption as these used small 0.5oz increments (equivalent to 2.9 cigarettes per day).

We calculated smoking intensity for current daily smokers only; smoking intensity for other participants was set to zero. As discussed in the previous section, it is worth noting that, in some sweeps, it was not possible to clearly identify occasional smokers, either through the available of a questionnaire item on smoking status or an clear or valid option to report smoking fewer than one cigarettes per day, on average. The potential inclusion of occasional smokers could bias average smoking intensity in the population downwards, though in practice, there appears to be relatively few occasional smokers in the data when they could be identified unambiguously.

We created smoking intensity variables for ages 20y, 25y, 31y, 36y, 43y, 53y, 63y, and 68y in the NSHD, ages 23y, 33y, 42y, 46y, 50y, and 55y in the NCDS, and ages 26y, 29y, 34y, 38y, 42y, and 46y in the BCS70. The restriction against categorical responses meant we did not use any data on smoking intensity from MCS participants. The variables and specific definitions used to define current smoking intensity are shown in Table 2 below.

Table 2: Variables used to define current smoking intensity (cigarettes per day) by cohort-sweep.

| **Cohort** | **Sweep** | **Definition** | **Variable(s)** | **Notes** |
| --- | --- | --- | --- | --- |
| BCS70 | 26y | Response to question on number of cigarettes usually smoked per day. | b960633 |  |
| BCS70 | 29y | Response to question on number of cigarettes usually smoked per day. | nofcigs |  |
| BCS70 | 34y | Response to question on number of cigarettes usually smoked per day. | b7nfcigs |  |
| BCS70 | 38y | Response to question on number of cigarettes usually smoked per day. | b8nfcigs |  |
| BCS70 | 42y | Response to question on number of cigarettes usually smoked per day. | b9nfcigs |  |
| BCS70 | 46y | Response to question on number of cigarettes usually smoked per day. | b10nfcigs |  |
| NCDS | 23y | Response to question on number of cigarettes usually smoked per day, with participant prompted to take average over a week if number varies. | n5935 |  |
| NCDS | 33y | Response to question on number of cigarettes usually smoked per day. | n504263 |  |
| NCDS | 42y | Response to question on number of cigarettes usually smoked per day. | nofcigs |  |
| NCDS | 46y | Response to question on number of cigarettes usually smoked per day. | n7nfcigs |  |
| NCDS | 50y | Response to question on number of cigarettes usually smoked per day. | n8nfcigs |  |
| NCDS | 55y | Response to question on number of cigarettes usually smoked per day. | n9nfcigs |  |
| NSHD | 20y | Derived from responses to questions on (a) usual number of manufactured cigarettes smoked per day on weekdays, and (b) ounces of rolling tobacco used per week. | SMODS66, SMOR66 |  |
| NSHD | 25y | Derived from responses to questions on (a) usual number of manufactured cigarettes smoked per day on weekdays, (b) usual number of manufactured cigarettes smoked per day on weekends, and (c) ounces of rolling tobacco used per week. | SMOD71, SMOW71, SMOR71 |  |
| NSHD | 31y | Response to question on usual number of cigarettes smoked per day over last year | SMODS77 |  |
| NSHD | 36y | Response to question on cigarettes smoked *now* per day | SMODS82 |  |
| NSHD | 43y | Response to question on cigarettes smoked *now* per day | SMODS89 |  |
| NSHD | 53y | Response to question on cigarettes smoked *now* per day | SMODS | Note on questionnaire asks to give equivalent number of cigarettes if roll-ups, but it isn’t clear if the nurse does this calculation or the cohort member. |
| NSHD | 63y | Response to question on cigarettes smoked *now* per day | SMOD09 | Cohort member asked to give equivalent number of cigarettes |
| NSHD | 68y | Response to question on cigarettes smoked *now* per day | SMOD14x | Cohort member asked to give equivalent number of cigarettes and explicitly asked to not include e-cigarettes |

#### Cohort Member Former Daily Smoking

We defined former regular smoking as smoking 1+ cigarettes per day for a period of 12 months or more (yes/no). Data on former regular smoking was not available in the MCS, but was available in multiple sweeps of the NSHD, NCDS, and BCS70. Thankfully in most of these sweeps, items on former smoking followed the definition used here – i.e., participants were direct asked if they had smoked 1+ cigarettes per day for a period of 12 months or more. Items in a small number of sweeps were worded less precisely (e.g., age 20y in the NSHD), either by not specifying smoking daily or clarifying the minimum period consumption over which consumption had to be maintained. To maintain consistency, we disregarded data from these sweeps.

We treated current daily smoking and former daily smoking as mutually exclusive; we classified all current daily smokers as non-former smokers. As noted, this could introduce inaccuracies given differences in the timeframe the current and former daily smoking variables refer to. We allowed current occasional smokers (< 1 cigarettes each day) to be classified as former smokers. However, this created inaccuracies in several sweeps where issues with questionnaire design meant occasional smokers were not routed into questions on former regular smoking (ages 20y, 36y, 53y, 63y, and 68y in the NSHD and age 23y in the NCDS). As there were over 200 occasional smokers at ages 20y in the NSHD and 23y in the NCDS, we did not create variables on former daily smoking for these sweeps. However, in later sweeps of the NSHD, there appeared to be far few occasional smokers, so we made former smoking variables for these sweeps, regardless.

We created former daily smoking variables at ages 25y, 36y, 43y, 53y, 63y and 68y in the NSHD, ages 33y, 42y, 46y and 50y in the NCDS, and ages 29y, 34y, 42y and 46y in the BCS70. Note, though being a former smoker is a permanent attribute, for each sweep, we only used responses on former smoking from that sweep to define former smoking (i.e., we did not feed-forward former smoking information from time *t* to define former smoking at time *t+1*). The variables and operationalizations used to define current smoking intensity are shown in Table 3 below.

Table 3: Variables used to define former daily smoking by cohort-sweep.

| **Cohort** | **Sweep** | **Definition** | **Variable(s)** | **Notes** |
| --- | --- | --- | --- | --- |
| BCS70 | 29y | Responds yes to question on ever smoking 1+ cigarettes a day for as long as a year | exsmoker |  |
| BCS70 | 34y | Responds yes to question on ever smoking 1+ cigarettes a day for as long as a year | b7exsmer |  |
| BCS70 | 42y | Responds yes to question on ever smoking 1+ cigarettes a day for as long as a year | B9EXSMER |  |
| BCS70 | 46y | Responds yes to question on ever smoking 1+ cigarettes a day for as long as a year | B10EXSMER |  |
| NCDS | 33y | Responds yes to question on ever smoking 1+ cigarettes a day for as long as a year | n504265 |  |
| NCDS | 42y | Responds yes to question on ever smoking 1+ cigarettes a day for as long as a year | exsmoker |  |
| NCDS | 46y | Responds yes to question on ever smoking 1+ cigarettes a day for as long as a year | n7exsmer |  |
| NCDS | 50y | Responds yes to question on ever smoking 1+ cigarettes a day for as long as a year | N8EXSMER |  |
| NSHD | 25y | Responds yes to question on ever smoking 1+ cigarettes a day or 1+ oz of rolling tobacco a month for as long as a year | SMO71 |  |
| NSHD | 36y | Responds yes to question on ever smoking 1+ cigarettes a day for as long as a year | SMOSE82 | Occasional smokers are in theory routed away from this question, but in the data there appear to be no occasional smokers (which may be due to occasional smokers not recording < 1 cigarettes per day). |
| NSHD | 43y | Responds yes to question on ever smoking 1+ cigarettes a day for as long as a year | SMOSE89 |  |
| NSHD | 53y | Responds yes to question on ever smoking 1+ cigarettes a day for as long as a year | SMOSE | Occasional smokers are in theory routed away from this question, but in the data there appear to be no occasional smokers (which may be due to occasional smokers not recording < 1 cigarettes per day). |
| NSHD | 63y | Responds yes to question on ever smoking 1+ cigarettes a day for as long as a year | SMOR09 | Occasional smokers are routed away from this question, but there are only 4 occasional smokers identifiable in the data (SMOD09 == 666) |
| NSHD | 68y | Responds yes to question on ever smoking 1+ cigarettes a day for as long as a year | SMOR14x | Occasional smokers are routed away from this question, but there is only 1 occasional smoker identifiable in the data (SMOD14X == 666) |

#### Cohort Member Ever Daily Smoking

For each sweep that we were able to create current daily smoking and former daily smoking variables, we also derived a variable for every daily smoking. A participant was classified as ever daily smoking if, for the current sweep, they were defined as a current daily or a former daily smoker. A participant was instead classified as never daily smoking if they were neither classified as a current daily smoker nor a former daily smoker. As with former daily smoking, we only used data from the current sweep to define this variable. We created ever daily smoking variables at ages 25y, 36y, 43y, 53y, 63y and 68y in the NSHD, ages 33y, 42y, 46y and 50y in the NCDS, and ages 29y, 34y, 42y and 46y in the BCS70. The variables and operationalizations used to define current daily smoking and former daily smoking can be viewed in Table 1 and Table 3 above.

#### Parental Smoking During CM’s Childhood

The measures of parental smoking behaviour during cohort member’s childhoods differed along several dimensions, including the respondent, time period considered, and the tobacco products enquired about. Given this broad lack of consistency, we adopted a relatively inclusive approach and analysed every sweep of data available. Where possible we defined parental smoking as smoking pipe or cigar or smoking 1+ cigarettes per day and prioritised prospective measurement from the parent themselves (rather than the cohort member or their main caregiver), but this was not the case for all cohorts. Notably the NSHD only contained items given to cohort members when they were age 53y (SMOP1) and which of their parents smoked cigarettes, cigars or a pipe when they lived with them as a child, rather than at a specific age.

Given gender differences in smoking prevalence, we examined maternal and paternal smoking separately. We used data on male and female caregivers, rather than biological parents, specifically. Given the cohorts do not align in the ages in which parental smoking was enquired about (and the NSHD retrospectively asks about smoking during CM’s childhood in general) we used every available measurement occasion.

The variables and operationalizations used to define parental smoking are shown in Table 4 below.

Table 4: Variables used to define parental smoking by cohort-sweep.

| **Cohort** | **Focal Person** | **Focal Period** | **Sweep** | **Definition** | **Variables** | **Notes** |
| --- | --- | --- | --- | --- | --- | --- |
| MCS | Mother / Father | Current | 9m | Smokes 1+ cigarettes a day or cigars or pipe | APSMUS0[A-D], APSMMA00 | Reported by parent themselves |
| MCS | Mother / Father | Current | 3y | Smokes 1+ cigarettes a day or cigars or pipe | BPSMUS0[A-F], APSMMA00 | Reported by parent themselves |
| MCS | Mother / Father | Current | 5y | Smokes 1+ cigarettes a day or cigars or pipe | CPSMUS0[A-D], CPSMMA00 | Reported by parent themselves |
| MCS | Mother / Father | Current | 7y | Smokes 1+ cigarettes a day or cigars or pipe | DPSMUS0[A-E], DPSMMA00 | Reported by parent themselves |
| MCS | Mother / Father | Current | 11y | Smokes 1+ cigarettes a day or cigars or pipe | EPSMUS0[A-I], EPSMMA00 | Reported by parent themselves |
| MCS | Mother / Father | Current | 14y | Smokes 1+ cigarettes a day or cigars or pipe | FPSMUS0[A-G], GPSMMA00 | Reported by parent themselves |
| BCS70 | Mother | Current | 5y | Smokes 1+ cigarettes a day or cigars or pipe | e258 | Reported by mother |
| BCS70 | Father | Current | 5y | Smokes 1+ cigarettes a day or cigars or pipe | e259 | Reported by mother |
| BCS70 | Mother | Current | 10y | Smokes 1+ cigarettes a day or cigars or pipe | e9_1, e9_2 | Reported by mother |
| BCS70 | Father | Current | 10y | Smokes 1+ cigarettes a day or cigars or pipe | e11_1, e11_2 | Reported by mother |
| BCS70 | Mother | Current | 16y | Responds ‘1. Yes, cigarettes’ or ‘2. Smokes cigars/cheroots/pipe’ to question on current smoking | og2_11 | Reported by mother |
| BCS70 | Mother’s Husband | Current | 16y | Responds ‘1. Yes, cigarettes’ or ‘2. Smokes cigars/cheroots/pipe’ to question on current smoking | og2_11 | Reported by mother. Father defined here as mother’s husband. |
| NCDS | Mother | Current | 16y | Respond ‘3. 1 to 5 per day’ or more or ‘pipe or cigar only’. | n2400 | Reported by main parent (typically mother) |
| NCDS | Father | Current | 16y | Respond ‘3. 1 to 5 per day’ or more or ‘pipe or cigar only’. | n2401 | Reported by main parent (typically mother) |
| NSHD | Parental Figures | Childhood | 53y | Respond yes to question on parents smoking cigarettes, cigars or a pipe when CM lived with parent as a child | SMOP1, SMOP2 | Reported retrospectively by cohort member. Note this does not separate cigar or pipe smoking though very few women smoke cigars or pipes (see NCDS 16y data). |

#### Maternal Smoking During Pregnancy with CM

Questions on maternal smoking during pregnancy were asked in the NCDS, BCS70 and MCS. Here we define maternal smoking during pregnancy as smoking daily. In the NCDS, smoking daily was defined as medium, heavy or variable smoking or medium or heavy smoking (sensitivity analysis) using the derived variable n639 – raw data are not available and it is unclear from the component questions or variables (n502 and n503) what variable refers to. In the BCS70, we also use a derived variable (a0043b). Maternal smoking is defined as continual smoking. In the MCS, we define smoking as continual smoking or as daily smoking during month 5 or after.

### Figures

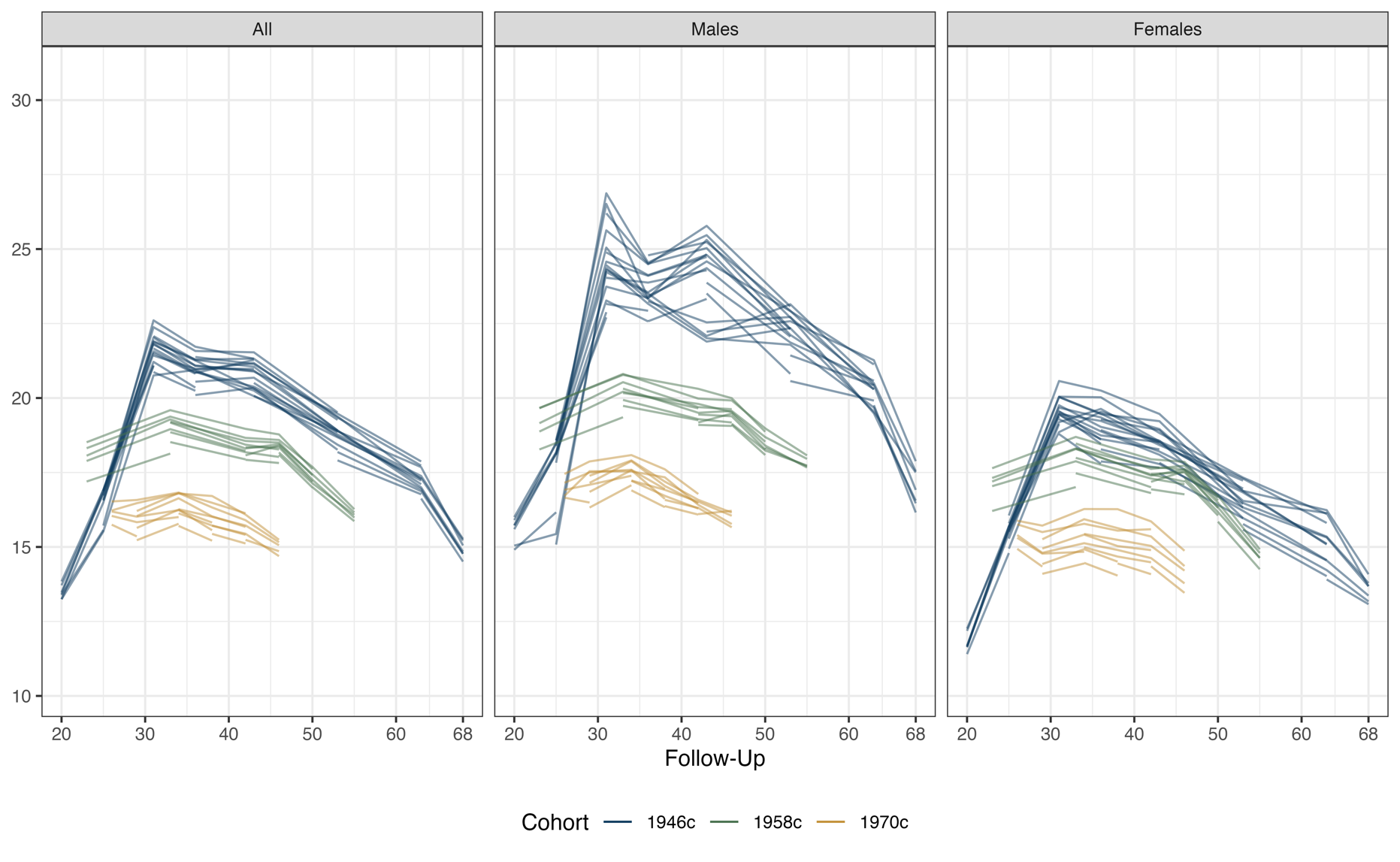

Figure S1: Average number of cigarettes smoked per day among sustained smokers. Each line represents a set of sustained smokers defined by reporting smoking daily at each sweep along the line (e.g., for the 1946c, the line spanning 20y-68y represents smokers reporting smoking at each sweep between and including 20y – 68y). Calculated using multiply imputed data (m = 40) and accounting for complex survey design.

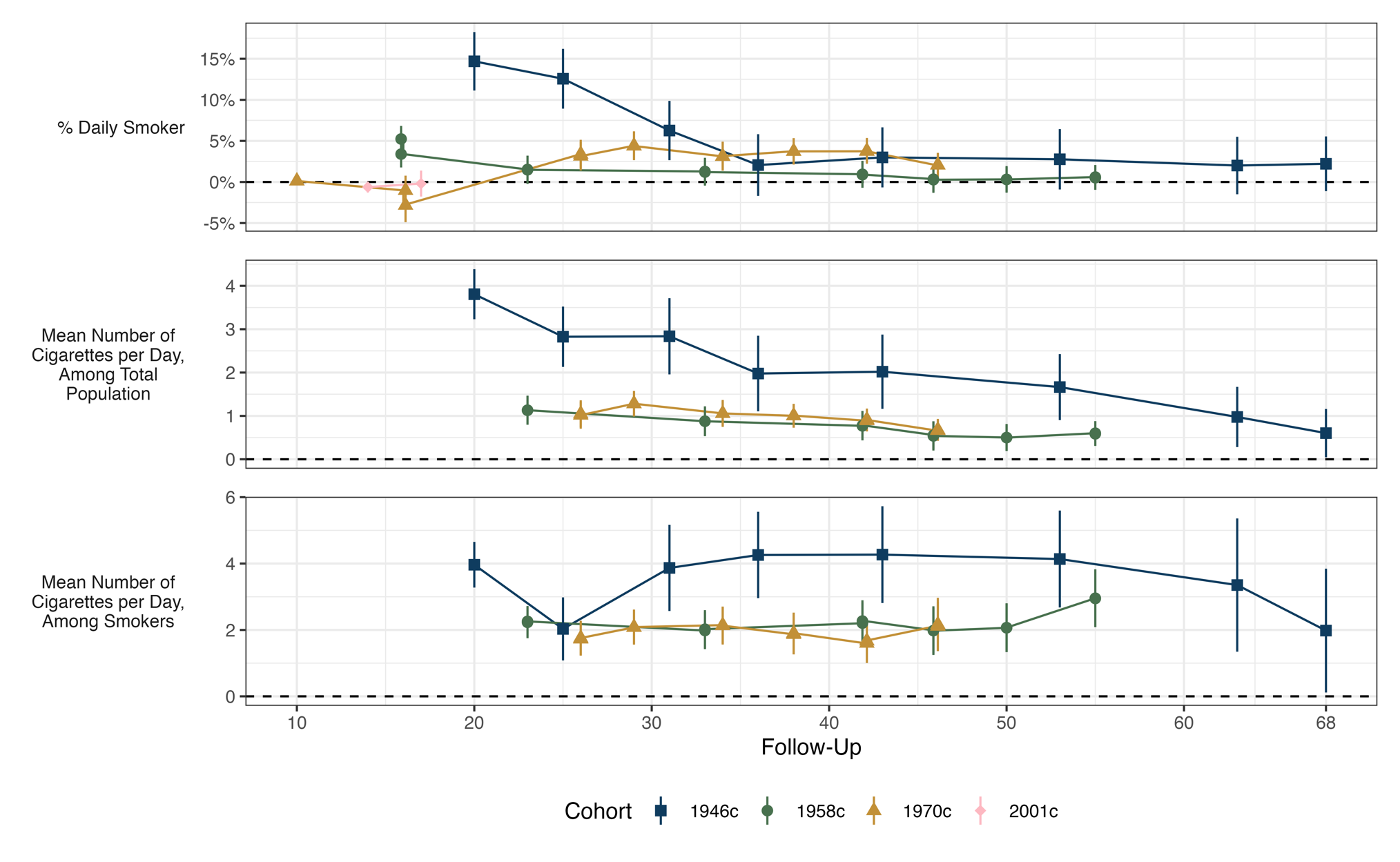

Figure S2: Sex-differences (male vs female) in cohort members’ smoking behaviour, by cohort and sweep. Values above zero represent great propensity of males to engage in the behaviour. Calculated using multiply imputed data (m = 40) and accounting for complex survey design

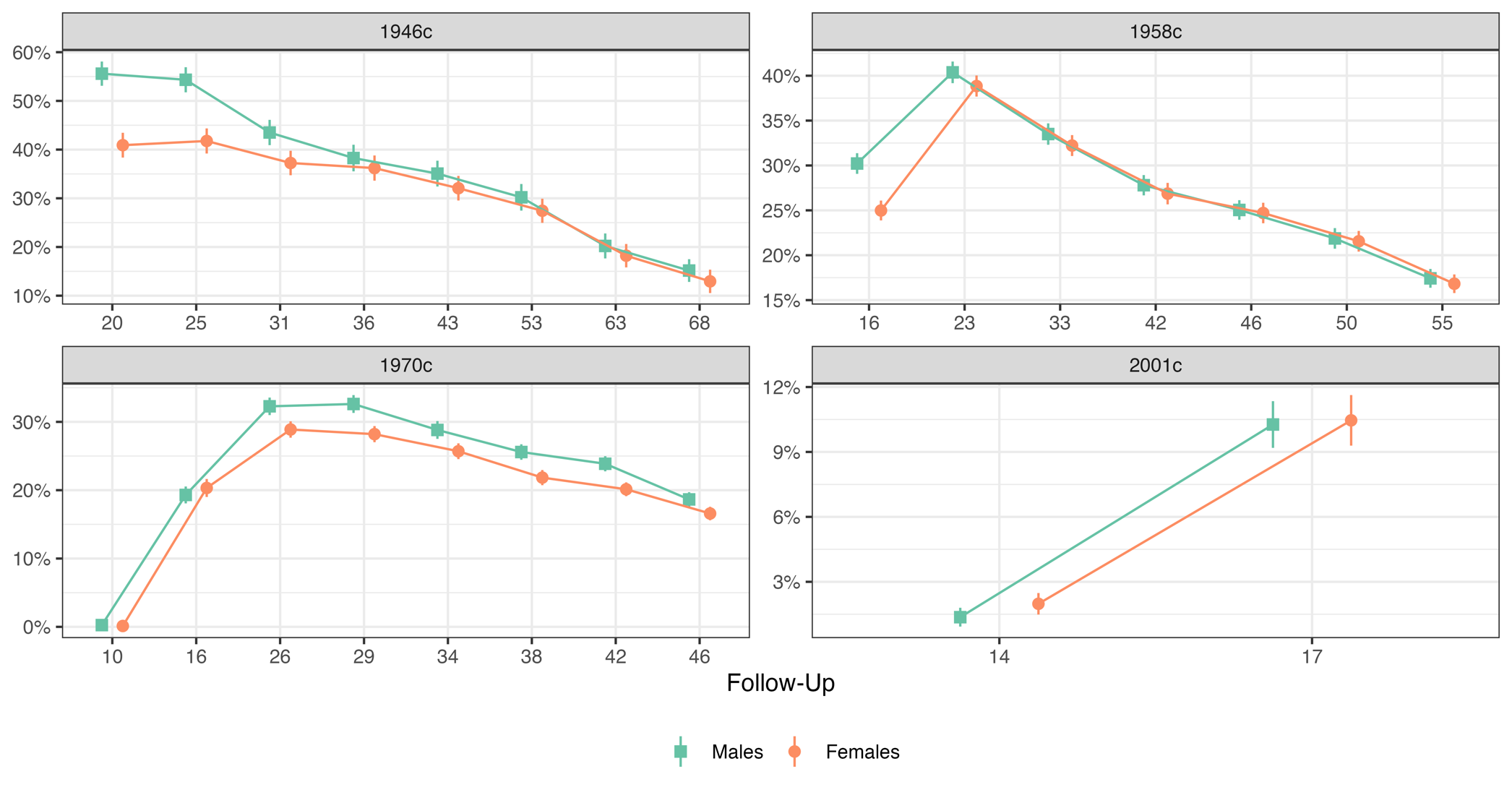

Figure S3: Percentage of cohort members who smoke daily, by cohort, sweep, and sex. Calculated using multiply imputed data (m = 40) and accounting for complex survey design.

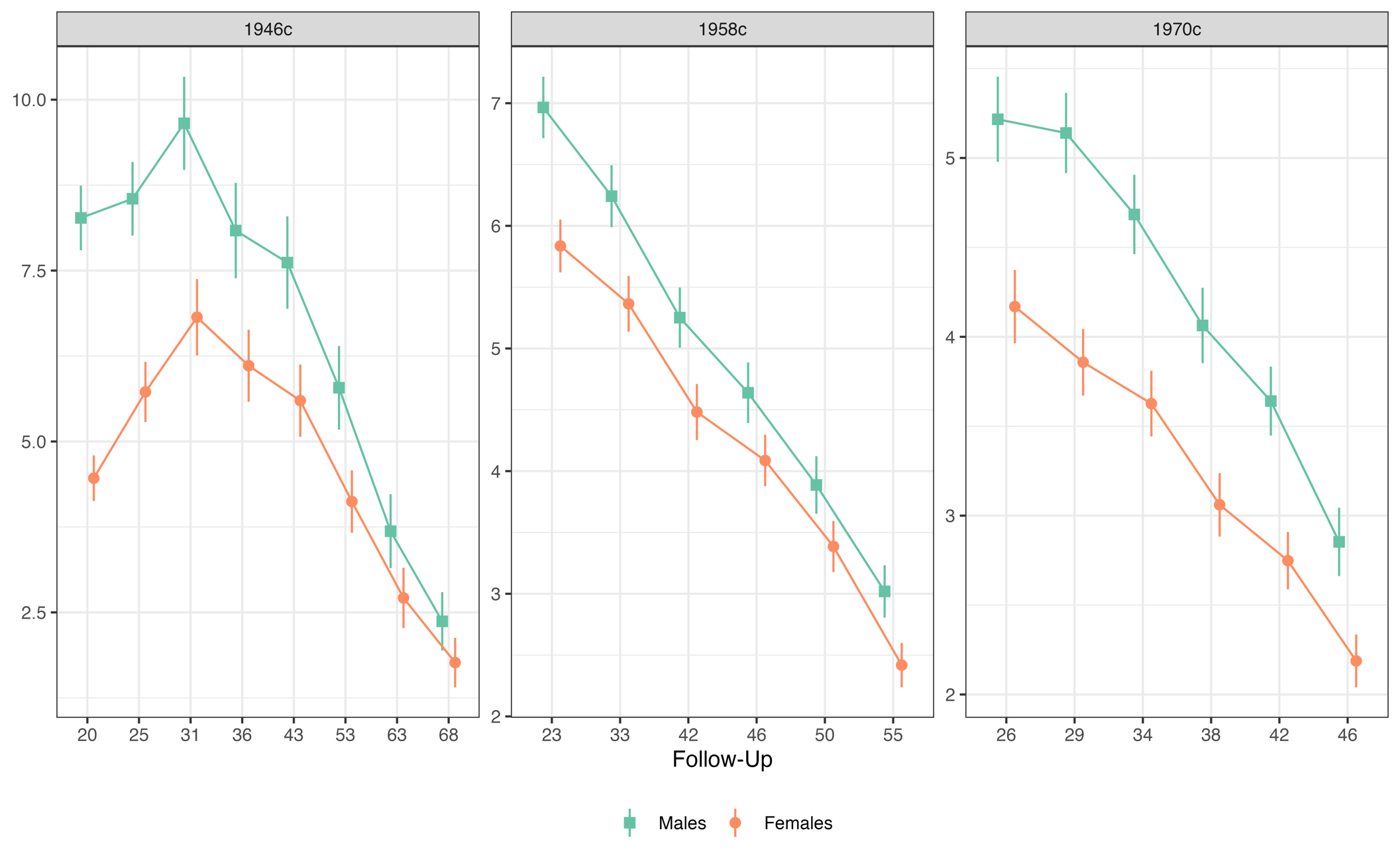

Figure S4: Mean number of cigarettes smoked per day among total sample by cohort, sweep, and sex. Calculated using multiply imputed data (m = 40) and accounting for complex survey design

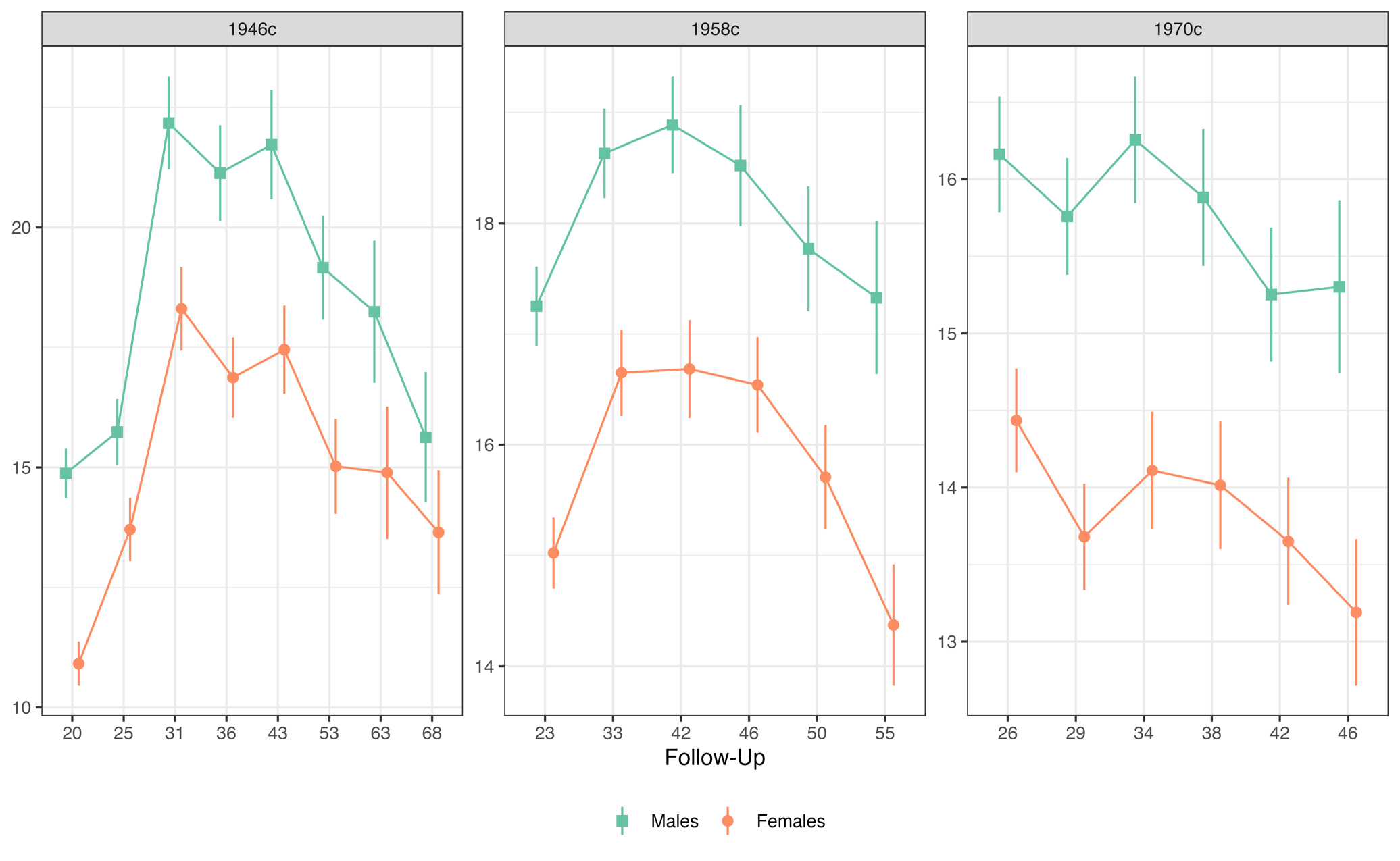

Figure S5: Mean number of cigarettes smoked per day among daily smokers by cohort, sweep, and sex. Calculated using multiply imputed data (m = 40) and accounting for complex survey design.

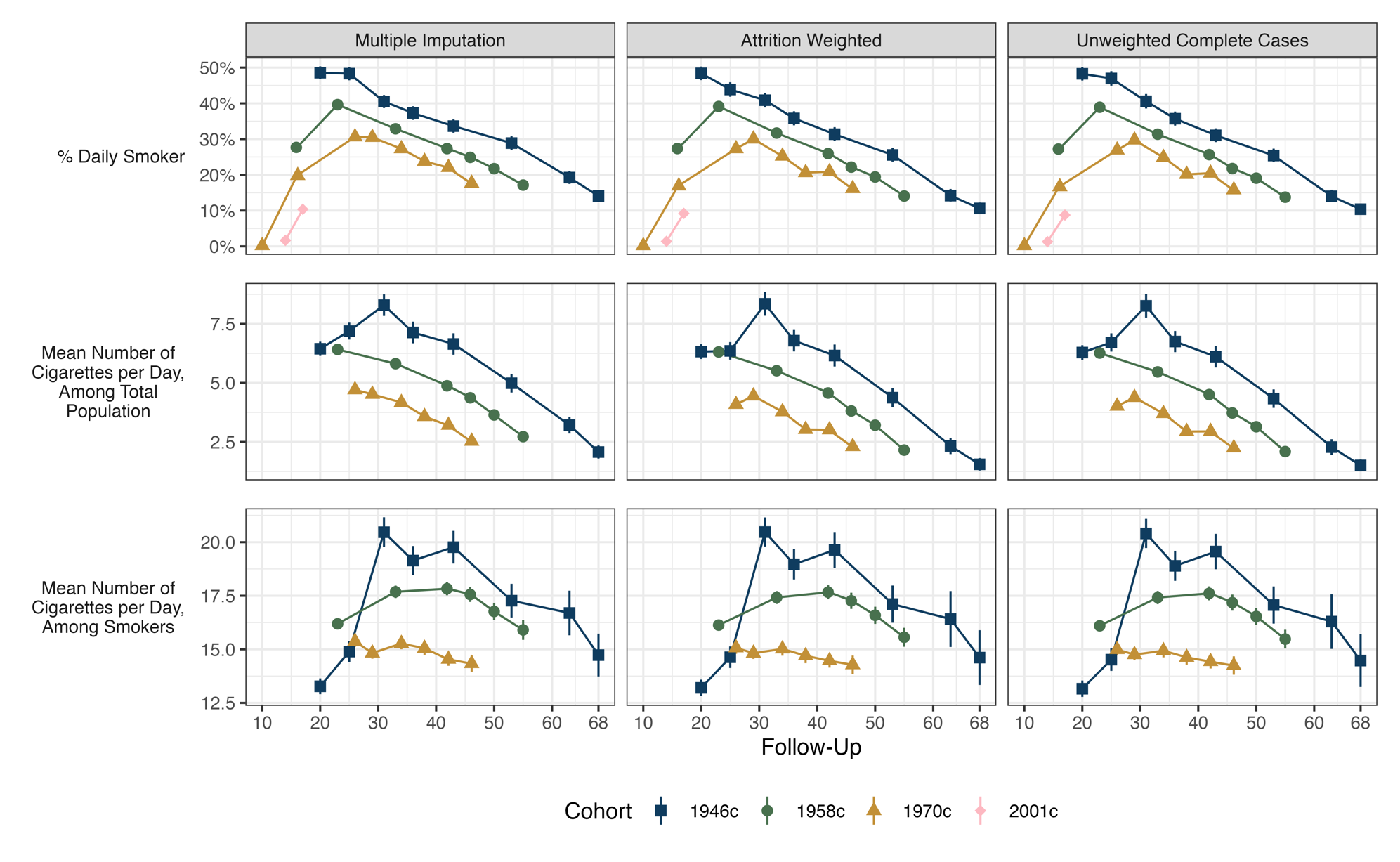

Figure S6: Cohort member smoking behaviour by cohort, sweep and estimator used. Left panel: multiple imputation. Middle panels: weighted for non-response. Right panels: complete case data, not weighted for non-response. All figures account for complex survey design.

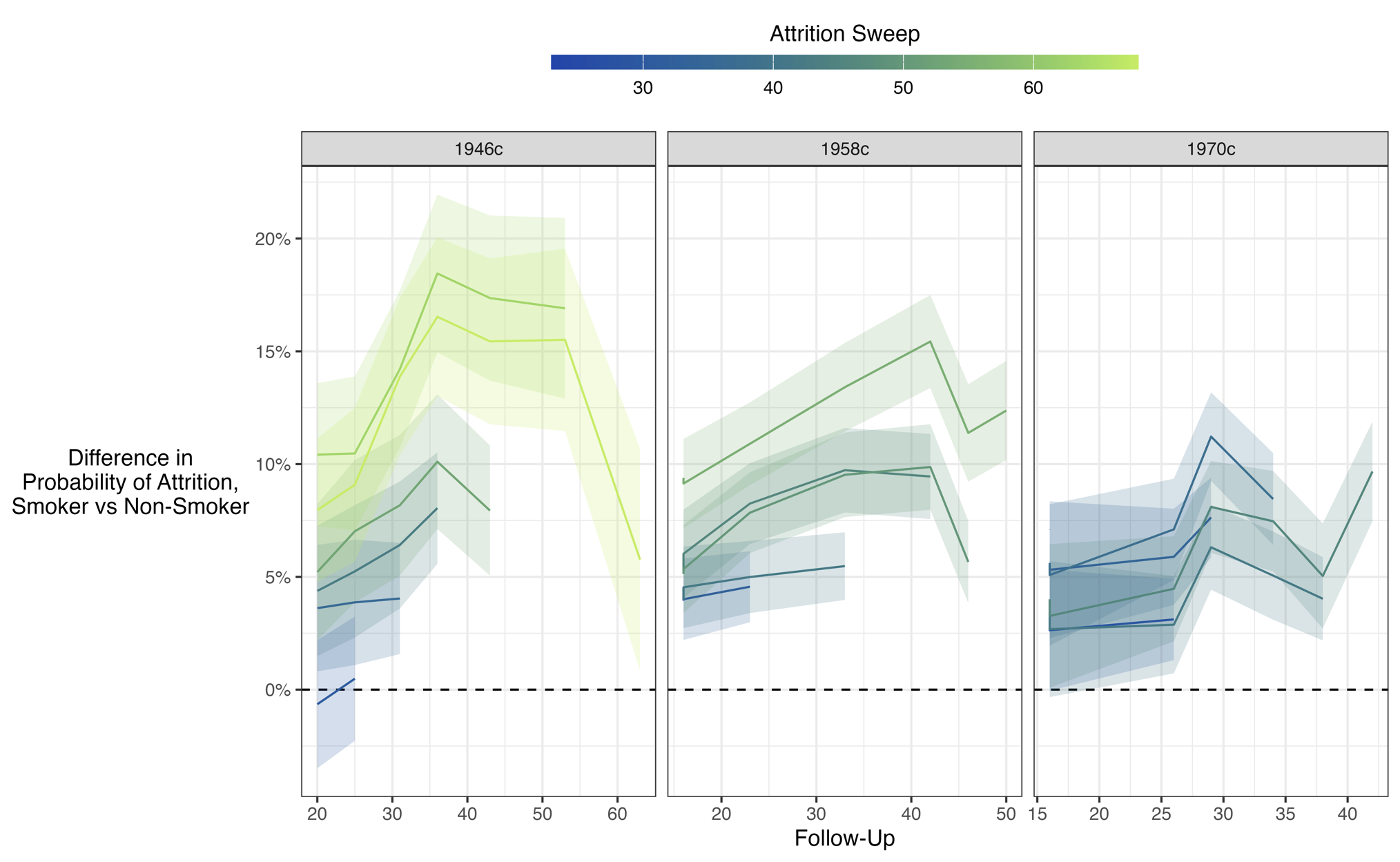

Figure S7: Difference in probability of not responding in a given sweep (indicated by line colour) among smokers vs. non-smokers by cohort and age at which smoking behaviour measured. Values above zero indicate smokers were less likely to respond at the sweep indicated by line coluor.

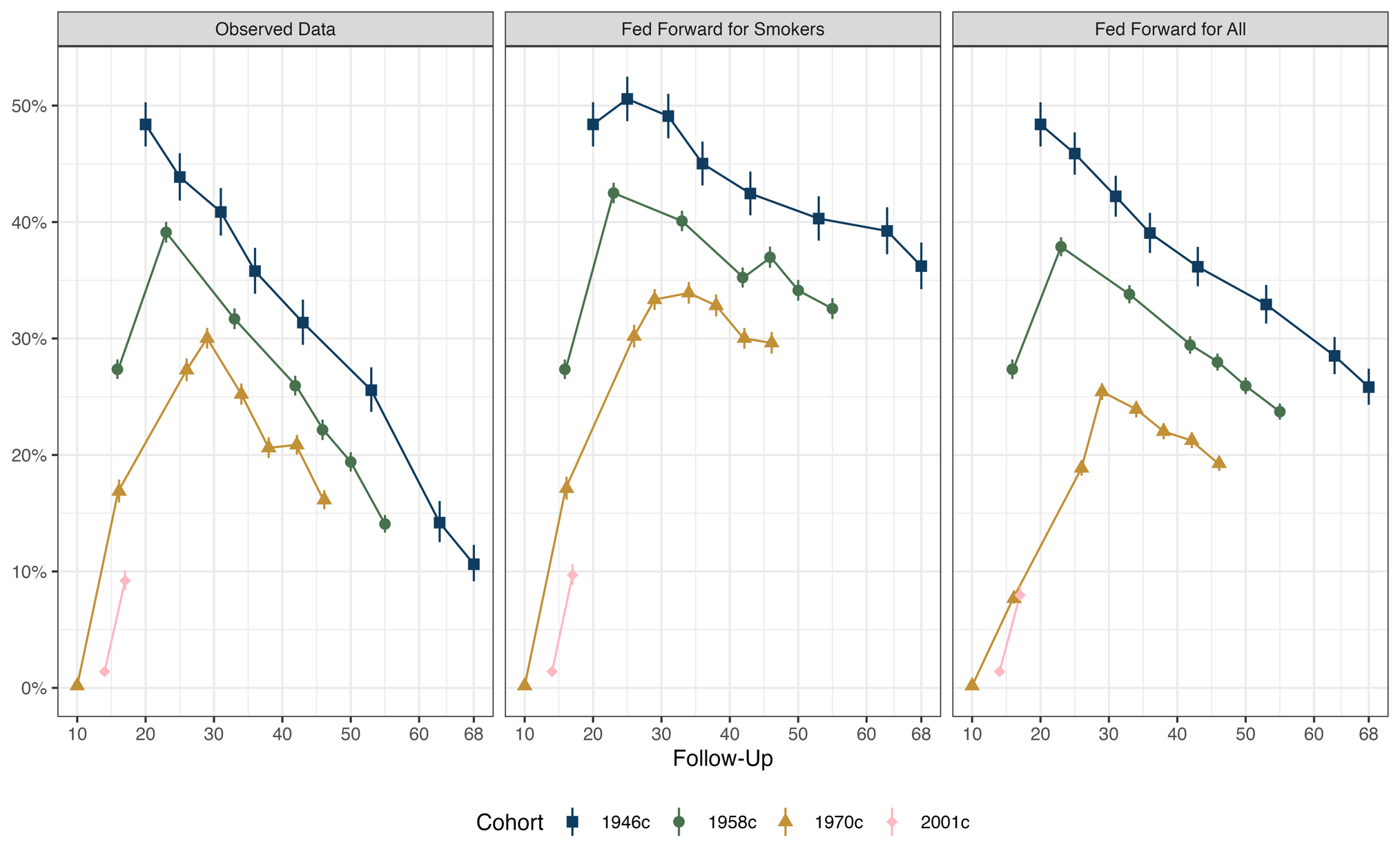

Figure S8: Estimated percentage of cohort members who smoke daily by cohort, sweep and whether previous daily smoking behaviour values were fed forward. Left panel: observed data (no feeding forward). Middle panel: feeding forward for smokers only (i.e., assumed to still be a smoker if missing and previous observed value was daily smoker). Right panel: feeding forward for all individuals (i.e., assumed to still be previous observed smoking status if missing for current sweep). All figures weighted for non-response and account for complex survey design.

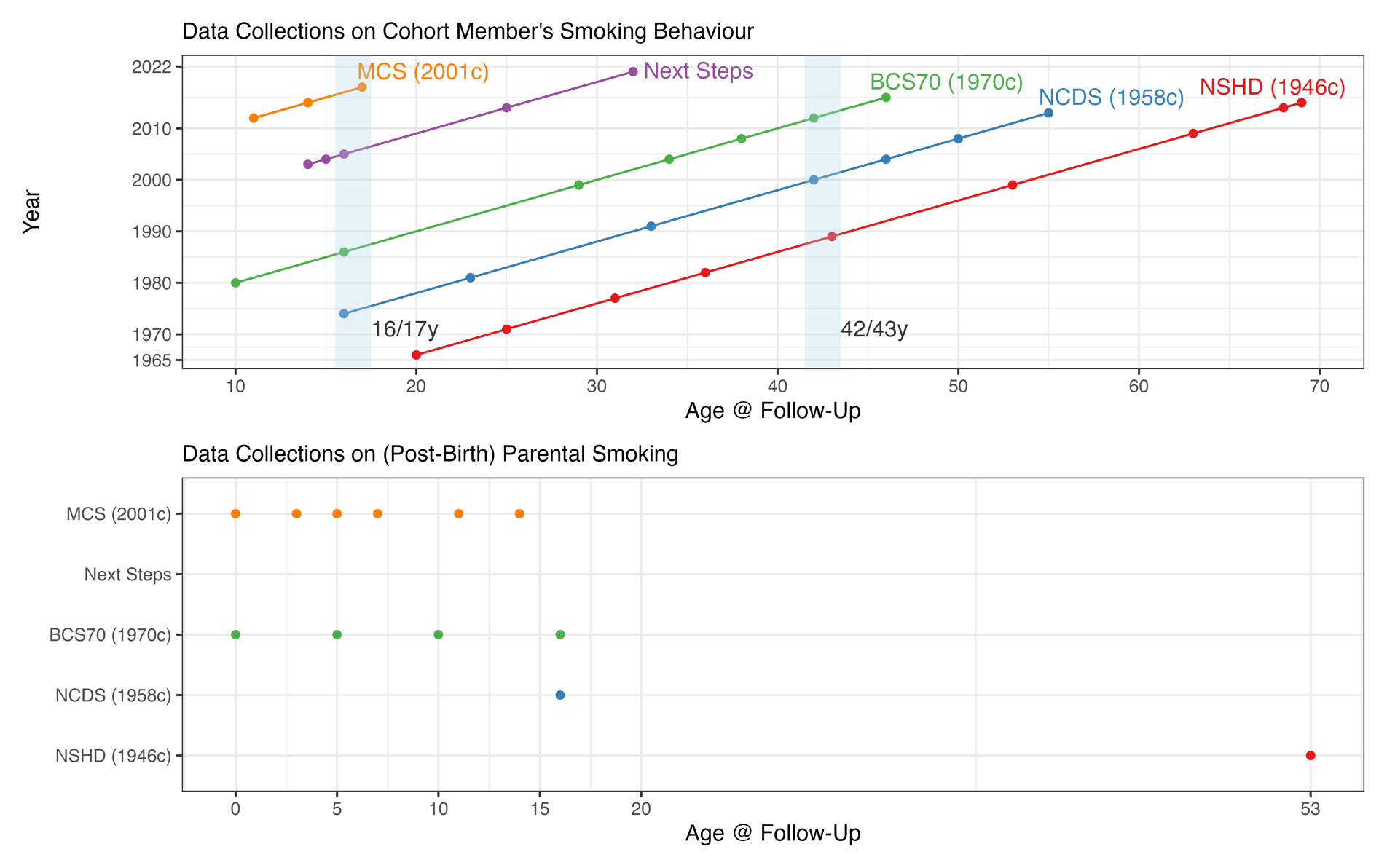

Figure S9: Data collection on smoking behaviour in the four British birth cohort studies and Next Steps. Blue shaded areas show age ranges in which (contemporary) smoking behaviour was collected in 3 or more cohorts.

1. Routing was used so this question was only answered by participants reporting ‘smoking at all nowadays’.
